# Impact of the COVID-19 Pandemic on anxiety and depression symptoms of young people in the Global South: evidence from a four-country cohort study

**DOI:** 10.1101/2021.02.02.21250897

**Authors:** Catherine Porter, Marta Favara, Annina Hittmeyer, Douglas Scott, Alan Sánchez Jiménez, Revathi Ellanki, Tassew Woldehanna, Le Thuc Duc, Michelle G. Craske, Alan Stein

**Author notes:** Correspondence to: Dr Catherine Porter, Lancaster University Management School, Bailrigg, Lancaster LA1 4YX, United Kingdom.

## Abstract

**Objective:** To provide evidence on the effect of the COVID-19 pandemic on the mental health of adolescents and young adults who grew up in poverty in Low and Middle Income Countries (LMICs).

**Design:** A phone survey implemented August-October 2020 to participants of a population-based cohort study since 2002 comprising two cohorts born in 1994-5 and 2001-2 in Ethiopia, India (Andhra Pradesh and Telangana), Peru and Vietnam. We examined associations between mental health and pandemic-related stressors, as well as structural factors (gender, location, wealth); and lifelong protective/risk factors (parent and peer relationship, past household wealth, long-term health problems, past emotional problems and subjective well-being) measured at younger ages.

**Setting:** A diverse, poverty focused sample, reaching those without mobile phones or internet access.

**Participants:** 10,496 individuals were approached, 9,730 participated. Overall, 8,988 individuals were included in this study, 4,610 (51%) male and 4,378 (49%) female. Non-inclusion was due to non-location or missing data.

**Main outcome measures:** At least mild anxiety and depression were measured by Generalized Anxiety Disorder-7 (GAD-7, ≥5) and Patient Health Questionnaire-8 (PHQ-8, ≥5).

**Results:** Rates of symptoms of at least mild anxiety (depression) were highest in Peru at 41% (32%) [95% CI, 38.63-43.12; (29.49-33.74)], and lowest in Vietnam at 9% (9%) [95% CI, 8.16-10.58; (8.33-10.77)], mirroring COVID-19 mortality rates. Females were most affected in all countries but Ethiopia. In all countries, pandemic-related stressors were associated with increased rates of anxiety and depression, though with varying levels of importance across countries. Prior parent and peer relationships were protective factors for mental health while having a long-term health problem or prior emotional problems were risk factors.

**Conclusion:** The COVID-19 pandemic presents significant risks to the mental health of young people. Mental health support is limited in LMICs and young people have to date been lower priority for COVID-19 interventions.

**Strengths and limitations of this study:**

- The study uses data from adolescents and young adults who grew up in poverty in four LMICs which were diversely affected by the COVID-19 pandemic, therefore investigating a globally vulnerable, but understudied group both in terms of age and wealth.
- This study reaches a broad sample of young people who grew up in poverty, including those without internet or mobile phone access.
- A key strength is combining a broad range of pandemic-related stressors from survey data on experiences of COVID-19 with previously measured information on longer-term risk and protective factors, therefore contributing to a more complete picture of COVID-19 effects.
- A limitation of the study is that it does not have a directly comparable pre-COVID baseline for depression/anxiety, however, proxy variables are used as a baseline and the explanatory variables capture dynamics that happened during the pandemic.
- A further limitation is possible underreporting due to stigma associated with mental health, despite piloting and validation, as well as possible bias in self-reported experiences of pandemic-related stressors due to feelings of anxiety or depression.

## INTRODUCTION

The Coronavirus disease 2019 (COVID-19) is creating concerns about the mental health of young people around the globe. There has been a call for research funders and researchers to “deploy resources to understand the psychological effects”^1^ of the COVID-19 pandemic and the ensuing “mental health crisis”.^2^ The crisis likely exacerbates previous risk factors of poverty and vulnerability. The Lancet Commission on Global Mental Health had already identified poverty as a key risk factor for the onset and persistence of mental disorders. ^3^ A recent study^4^ found that those with the lowest income were much more likely to suffer from anxiety and depressive disorders than their wealthier counterparts and point to the bidirectional causal relationship between poverty and mental health.

Several studies have examined the mental health impacts of the pandemic, predominantly in high-income countries.^5,6,7^ The few studies from Low-and-Middle-income countries (LMICs) have primarily relied on convenience samples, and internet-based surveys (e.g.^8-11^), unlikely reaching the rural poor, though one study^12^ investigated the effect of immediate lockdown orders on (adult) women’s mental health and experiences of intimate partner violence using a phone survey in rural Bangladesh.

Half of all mental health conditions develop by 14 years of age and 75% by early adulthood.^3^ In developed countries young women aged 16-24 are the most likely to have experienced a deterioration in mental health during the pandemic.^12^ Thus, understanding risk and protective factors during the pandemic at this age is critical to prevention, especially for the poorest. There is little research on the mental health of adolescents in LMICs, though they make up the bulk of the global adolescent population.^13^

This study examines the impact of the COVID-19 pandemic on the mental health of nearly 10,000 young people from a 20-year cohort study operating in four LMICs: Ethiopia, India (Andhra Pradesh and Telangana), Peru and Vietnam. When the cohorts were originally recruited, the objective was to ensure that families living in poverty were substantially represented. ^14-17^ Nowadays, these countries represent a diverse set of experiences during the pandemic, in terms of number and severity of cases as well as policy responses. Figure 1 shows that COVID-19 has had by far the most striking impact in Peru in terms of deaths per population, followed by India. In contrast, Vietnam has been hailed as a success story in controlling the spread of the virus.

**Figure 1.**
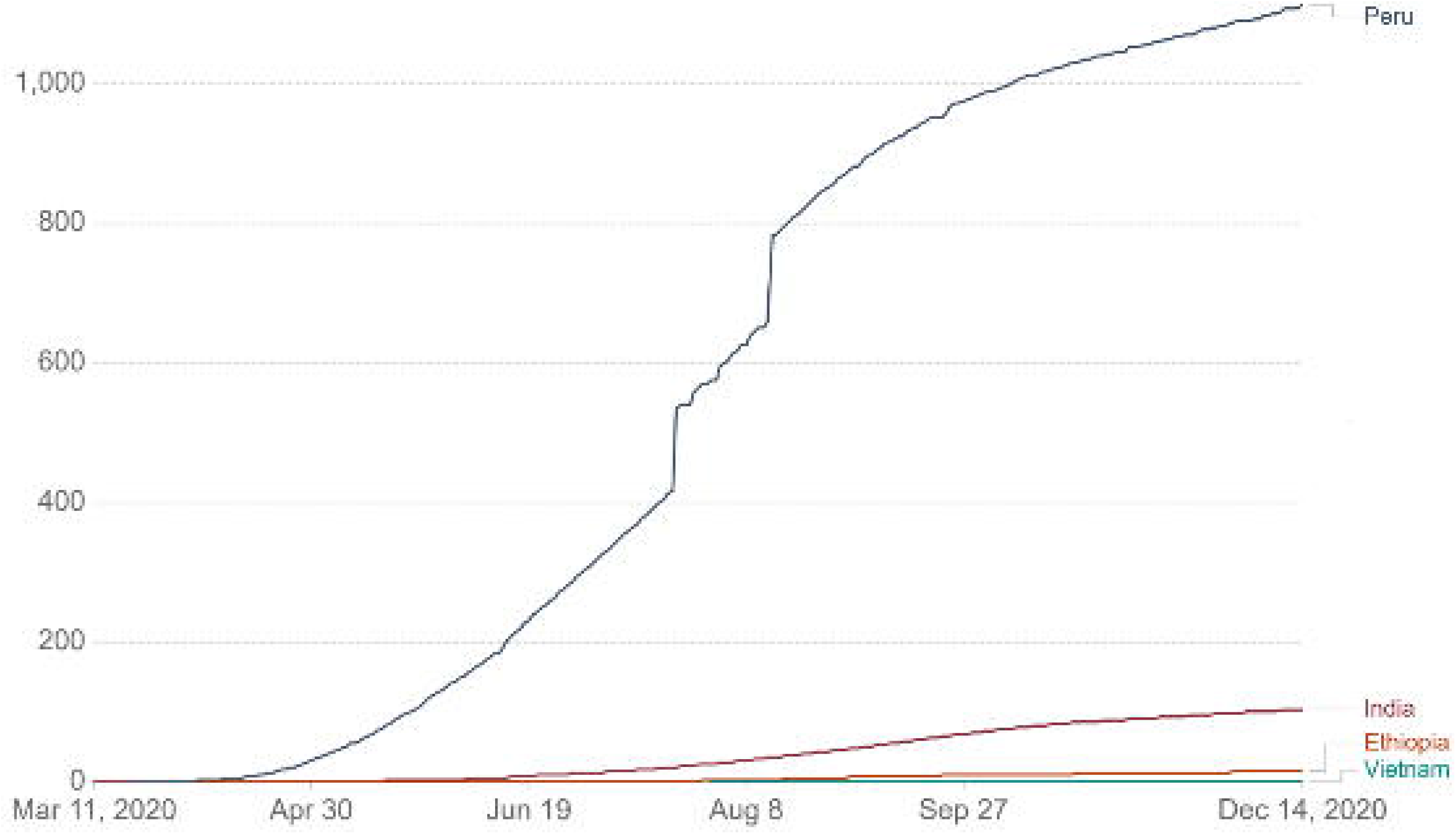
Cumulative confirmed COVID-19 cases per million people in the four Young Lives countries. Source: Johns Hopkins University CSSE COVID-19 Data^52^, accessed via our world in data^53^. Last updated 15 December 2020. Testing and challenges in the attribution of the cause of death means that the number of confirmed deaths may not be an accurate count of the true number of deaths from COVID-19.

Pressure on mental health is likely to be greater in countries which are more affected by the pandemic. Within each country, hypothesised stressors related to changes in circumstances/behaviours/wellbeing that occurred due to the pandemic (*COVID-19 related stressors*) include individuals’ perceived infection risk, economic adversities, changes in employment status and increased household responsibilities, educational disruption, and changes in subjective well-being between 2016 and the pandemic. Regarding *structural factors* (age, gender, rural/urban place of residence) we expect that residents of urban areas may have difficulties social distancing in slum-like conditions and possibly more likely than residents in rural areas to develop mental health conditions. Similarly, depression and anxiety symptoms might be more frequent among females than males. Changes imposed by COVID-19 in time use, education and work may impact the genders differently. We exploited information collected in previous survey rounds to investigate past *protective/risk factors measured during childhood and adolescence*: positive social interactions (parent and peer relations), past household wealth, and long-term health conditions. It also allows us to include *proxy baseline information for mental health* in the form of past emotional problems and subjective well-being measured 10 years ago at age 15.

## METHODS

### Study design and participants

A phone survey^18-21^ was administered between August and October 2020 to participants of the Young Lives study, a longitudinal survey established in 2002 following two cohorts of children born in 1994-5 and 2000-1 in Ethiopia, India (Andhra Pradesh and Telangana), Peru and Vietnam. Respondents have been surveyed in person every three years, with five consecutive rounds completed by 2016. In 2020, the cohort members are aged 18-19 (younger cohort) and 25-26 years (older cohort). The original sample was selected to include a significant coverage of poorer areas.^14-17^ 93% of the cohort (9,704) were tracked in 2019. The sample reduced to 8,988 individuals due to missing values for any question (Supplementary Table 1 and Supplementary Table 2) including between 0.1% (Vietnam) and 2% (Peru) who did not respond to the mental health questions (Supplementary Table 3).

The phone survey was administered over mobile telephone by up to 15 trained interviewers per country with access to hardware, software and internet access required for working from home. Responses were recorded in an electronic questionnaire using *Surveybe* Implementer software.

Symptoms of anxiety and depression were measured using the Generalized Anxiety Disorder-7 (GAD-7) scale and the Patient Health Questionnaire depression scale-8 (PHQ-8). The GAD-7 has been validated^22^ and used in all four study countries.^23-28^ The PHQ-9 was also validated^29-33^ and used in several studies.^25-28 34-39^ The scales were slightly adapted for administration in a phone survey. First, we asked participants whether they were alone in the room and if not, whether they could find a quiet space and/or make sure their phone speaker was off. Second, for each item in GAD-7 and PHQ-8 we asked whether the symptom had been observed (Yes/No) over the past 14 days, and if “Yes” we then asked about the frequency. The scales were administered as the last section of the survey.

GAD-7 scores between 5 and 9, 10 and 14 and above 15 represent mild, moderate, and severe anxiety, respectively.^40^ A PHQ-8 score between 5 and 9, 10 and 14, 15 and 19 and above 19 were considered representative of mild, moderate, moderately severe, and severe depression, respectively.^41^ Cronbach’s Alpha^42^ for both scales was close to or above 0.7.^43^ Inter-item correlations fell within the recommended range (0·15 to 0·50^44 45^; Supplementary Table 4).

### Statistical analysis

Logistic regressions were used to examine the relationship between a range of stressors on a binary variable indicating (at least) mild anxiety (GAD-7≥5) and (at least) mild depression (PHQ-8 ≥5), as reported in the Results section. We include four sets of stressors hypothesised to be associated with mental health: *changes in circumstances/behaviours/wellbeing that occurred due to the pandemic, past risk/protective factors, structural factors* and *proxy baseline information* (emotional problems and subjective wellbeing measured 10 years earlier). The characteristics of the sample population are shown in Supplementary Table 5. Figure 2 gives an overview of the variables used in the analysis and the respective ages when they were measured.

**Figure 2.**
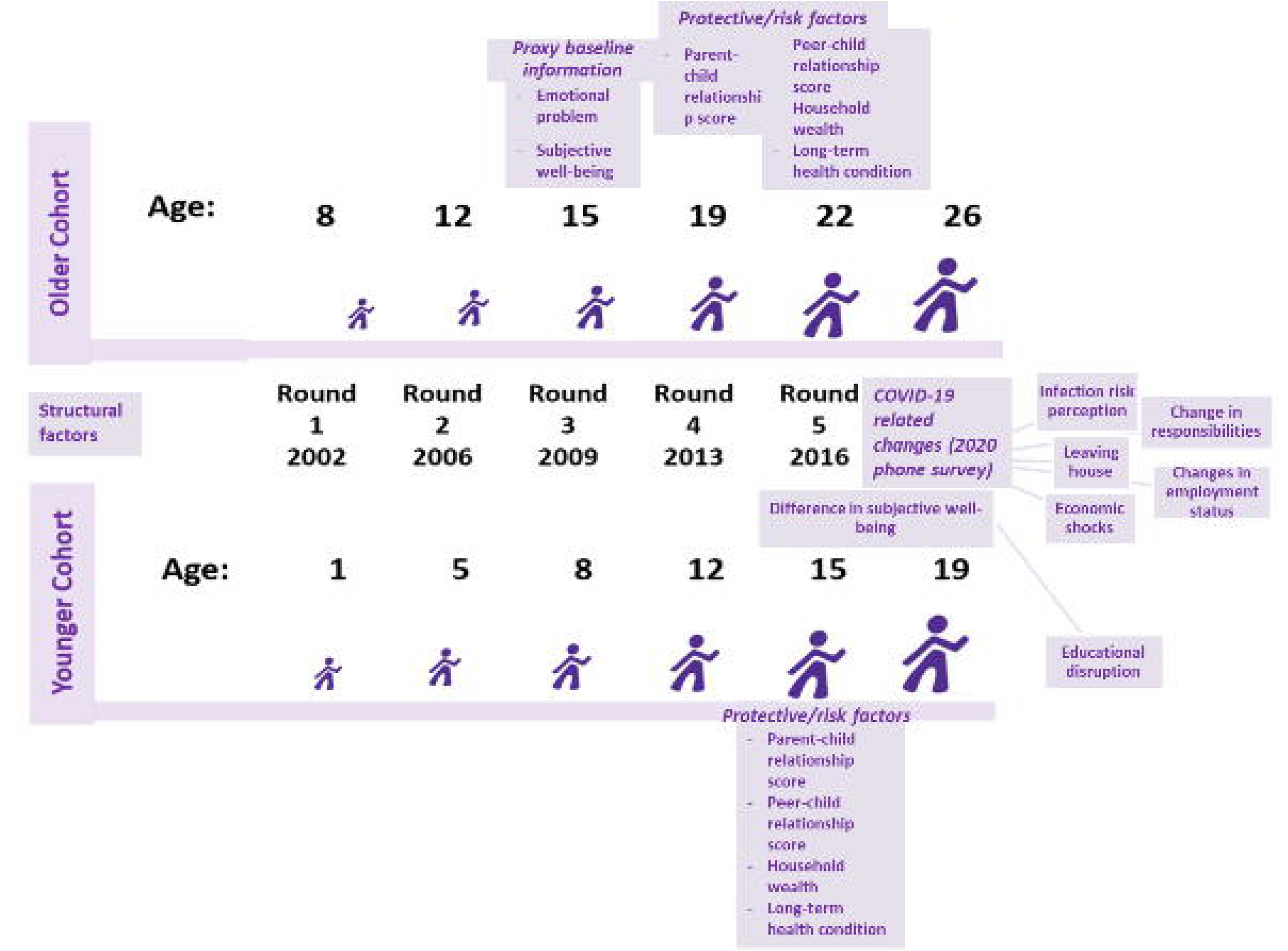
Variables used in analysis. Variables used in the analysis. Structural factors refer to age, gender and rural/urban location.

The first set of *COVID-19 related stressors* include perceived COVID-19 infection risk, the extent to which people practice self-isolation (having left the house in the past 7 days), increased household responsibilities (including spending more time caring for children, on household chores or working in a family business), suffering from any adverse economic events (including increases in the price of food, incurring increased health expenditures, fewer clients in a family business, and if so, whether the household reduced food consumption to cope with it) and changes in working status compared to before the pandemic. In further analysis for the 19-year-old cohort we replaced working status with engagement with education, given that more than half were still enrolled when the pandemic began (Supplementary tables 6 and 7). Finally, among the *COVID-19 related stressors*, we included the change in subjective well-being (SWB) between 2016 and the pandemic. SWB was measured in Round 5 (2016) at ages 15 and 22, and in the phone survey. Cantril’s Ladder (1965)^46^ asks respondents to visualise a ladder of nine steps; the bottom (top) step representing their worst (best) possible life. Respondents are asked to identify which step they presently stand on. Difference *in subjective well-being* is a continuous variable ranging from −8 to +8.

*Past risk and protective factors* include long-term health problems and the past household wealth^47^, both measured in 2016, and Parent-child and Peer-child relationships measured using the total raw scores of the Marsh Self-Description Questionnaires II^48^ and I^49^. Both scores range between 8 and 32, with higher scores being positive. Peer relationships were obtained at ages 15 (younger cohort) and 22 (older cohort) in 2016. Parent relationship was obtained at ages 15 (younger cohort, 2016) and 19 (older cohort, 2013).

GAD-7 and PHQ-8 were not measured in previous survey rounds. Therefore, we control for p*roxy baseline information* including emotional problems and SWB, both available for the 25-year-old cohort only in 2009 (round 3) at the age of 15 (Supplementary tables 8 and 9). The Emotional Problem Scale comes from the self-completed Strengths and Difficulties Questionnaire (SDQ). Total scores range from 0 to 10, a higher score indicates more severe emotional problems. We report extensive disaggregated rates of mental health issues (Supplementary table 10 to supplementary table 15).

Changes in responsibilities, the labour market and education environment may affect males and females differently. Therefore, we re-estimated the regressions separately by gender (Supplementary table 16 to supplementary table 23). We report odds ratios, robust standard errors and the 95% confidence intervals for all regressions.

### Ethics statement

The survey was approved by the institutional research ethics committees at the University of Oxford (UK), the University of Addis Ababa (Ethiopia), the Centre for Economic and Social Studies in Hyderabad (India), the Instituto de Investigación Nutricional (Peru) and the Hanoi University of Public Health (Vietnam). Participants were asked for their verbal informed consent before the study commenced and were assured of confidentiality. A consultation guide was provided to all participants with resources for support in issues raised by the questionnaire, including mental health.

### Patient and public involvement

No patients or the public were involved in the study design, setting the research questions, interpretation or writing up of results, or reporting of the research.

## RESULTS

93% of the Young Lives sample located during the last tracking exercise in November 2019 across the four countries participated in the phone survey and only between 0·1% (Vietnam) and 2% (Peru) did not respond to the mental health questions. The results presented in this section refer to the 19-year olds and 25-year olds together, unless differently specified.

Both mild anxiety (41%, 95% CI 38·63-43·12) and mild depression (32%, 29·49-33·74) rates were highest in Peru, followed by Ethiopia (anxiety: 18%, 16·28-19·54; depression: 15%, 13·95-17·02), see Table 1. The rates of moderate/severe anxiety and depression were highest in Peru, 13·5% (95% CI 12·00-15·14) and 9·6% (8·35-11·07), and below 3% in the other countries (Supplementary tables 24 to 31). Females had significantly higher rates of anxiety symptoms in all countries except Ethiopia and higher rates of depression in Peru and Vietnam. In Peru, almost half of all females had symptoms consistent with at least mild anxiety. Rates of anxiety and depression in rural areas were significantly lower than urban rates in Ethiopia and Peru, but significantly higher in India. The poorest wealth tercile had significantly lower rates of anxiety in Ethiopia and Peru, but higher rates in India and Vietnam. In Peru, the poorest wealth tercile also had significantly lower rates of depression. Those not having any access to the internet, although a minority, had significantly higher levels of anxiety (Vietnam p<0·01, Peru p<0·1).

**Table 1.** Rates of at least mild depression and anxiety in Ethiopia, India, Peru and Vietnam.

| Rates of at least mild anxiety | Ethiopia |  | India |  | Peru |  | Vietnam |  |
| --- | --- | --- | --- | --- | --- | --- | --- | --- |
|  | % at least mild Anxiety | CI95% | % at least mild Anxiety | CI95% | % at least mild Anxiety | CI95% | % at least mild Anxiety | CI95% |
| **Total** | **17.87** | **16.28; 19.54** | **11.06** | **9.88; 12.32** | **40.86** | **38.63; 43.12** | **9.32** | **8.16; 10.58** |
| Male | 17.48 | 15.30; 19.65 | 9.28*** | 7.74; 10.81 | 33.65*** | 30.65; 36.64 | 7.92** | 6.33; 9.51 |
| Female | 18.32 | 15.93; 20.70 | 13.01 | 11.14; 14.87 | 48.28 | 45.06; 51.49 | 10.63 | 8.88; 12.39 |
| Rural | 14.67*** | 12.65; 16.70 | 11.81* | 10.34; 13.27 | 33.71*** | 28.79; 38.63 | 9.07 | 7.47; 10.67 |
| Urban | 21.61 | 19.06; 24.16 | 9.20 | 7.13; 11.27 | 42.52 | 40.04; 45 | 9.62 | 7.83; 11.40 |
| Poorest tercile | 14.90** | 12.29; 17.51 | 13.52*** | 11.26; 15.78 | 36.92** | 33.01; 40.84 | 11.67*** | 9.41; 13.92 |
| Middle/Richest terciles | 19.32 | 17.29; 21.34 | 9.82 | 8.42; 11.21 | 42.63 | 39.94; 45.32 | 8.11 | 6.74; 9.49 |
| No internet | 15.97** | 13.82; 18.11 | 12.16 | 8.43; 15.89 | 51.32* | 40; 62.63 | 22.86*** | 8.74; 36.98 |
| With internet | 19.87 | 17.47; 22.27 | 10.92 | 9.65; 12.19 | 40.42 | 38.16; 42.68 | 9.11 | 7.92; 10.30 |

| Rates of at least mild depression | Ethiopia |  | India |  | Peru |  | Vietnam |  |
| --- | --- | --- | --- | --- | --- | --- | --- | --- |
|  | % at least mild Depression | CI95% | % at least mild Depression | CI95% | % at least mild Depression | CI95% | % at least mild Depression | CI95% |
| **Total** | **15.44** | **13.95; 17.02** | **9.92** | **8.80; 11.12** | **31.58** | **29.49; 33.74** | **9.49** | **8.33; 10.77** |
| Male | 14.92 | 12.88; 16.96 | 9.64 | 8.08; 11.21 | 27.38*** | 24.55; 30.21 | 7.65*** | 6.09; 9.22 |
| Female | 16.04 | 13.77; 18.31 | 10.22 | 8.54; 11.89 | 35.91 | 32.83; 39.00 | 11.22 | 9.42; 13.02 |
| Rural | 12.89*** | 10.98; 14.81 | 10.84** | 9.43; 12.25 | 26.40** | 21.82; 30.99 | 9.39 | 7.77; 11.01 |
| Urban | 18.43 | 16.03; 20.83 | 7.60 | 5.70; 9.50 | 32.79 | 30.44; 35.14 | 9.62 | 7.83; 11.40 |
| Poorest tercile | 14.35 | 11.78; 16.91 | 10.80 | 8.74; 12.85 | 26.15*** | 22.59; 29.72 | 8.46 | 6.51; 10.42 |
| Middle/Richest terciles | 15.97 | 14.10; 17.85 | 9.47 | 8.10; 10.85 | 34.02 | 31.45; 36.60 | 10.03 | 8.51; 11.54 |
| No internet | 14.09* | 12.06; 16.13 | 10.14 | 6.69; 13.58 | 31.58 | 21.05; 42.11 | 14.29 | 2.52; 26.05 |
| With internet | 16.85 | 14.60; 19.11 | 9.89 | 8.67; 11.10 | 31.58 | 29.44; 33.73 | 9.42 | 8.22; 10.63 |
| N | 2183 |  | 2622 |  | 1887 |  | 2296 |  |

Notes: If any missing answers to questions then the whole score is set to missing. \*\*\* $p < 0.01$ ; \*\* $p < 0.05$ ; \* $p < 0.1$ . Stars represent significance of t-test of equality between groups (Male-Female; Rural-Urban; Bottom-Top/Middle wealth tercile; Internet access through home computer/working smartphone (No-Yes)). Poorest and middle/richest terciles refer to the household's position in the 2016 (round 5) wealth distribution. Internet access refers to having internet access through a home computer and/or a working smart phone.

**Table 2.**
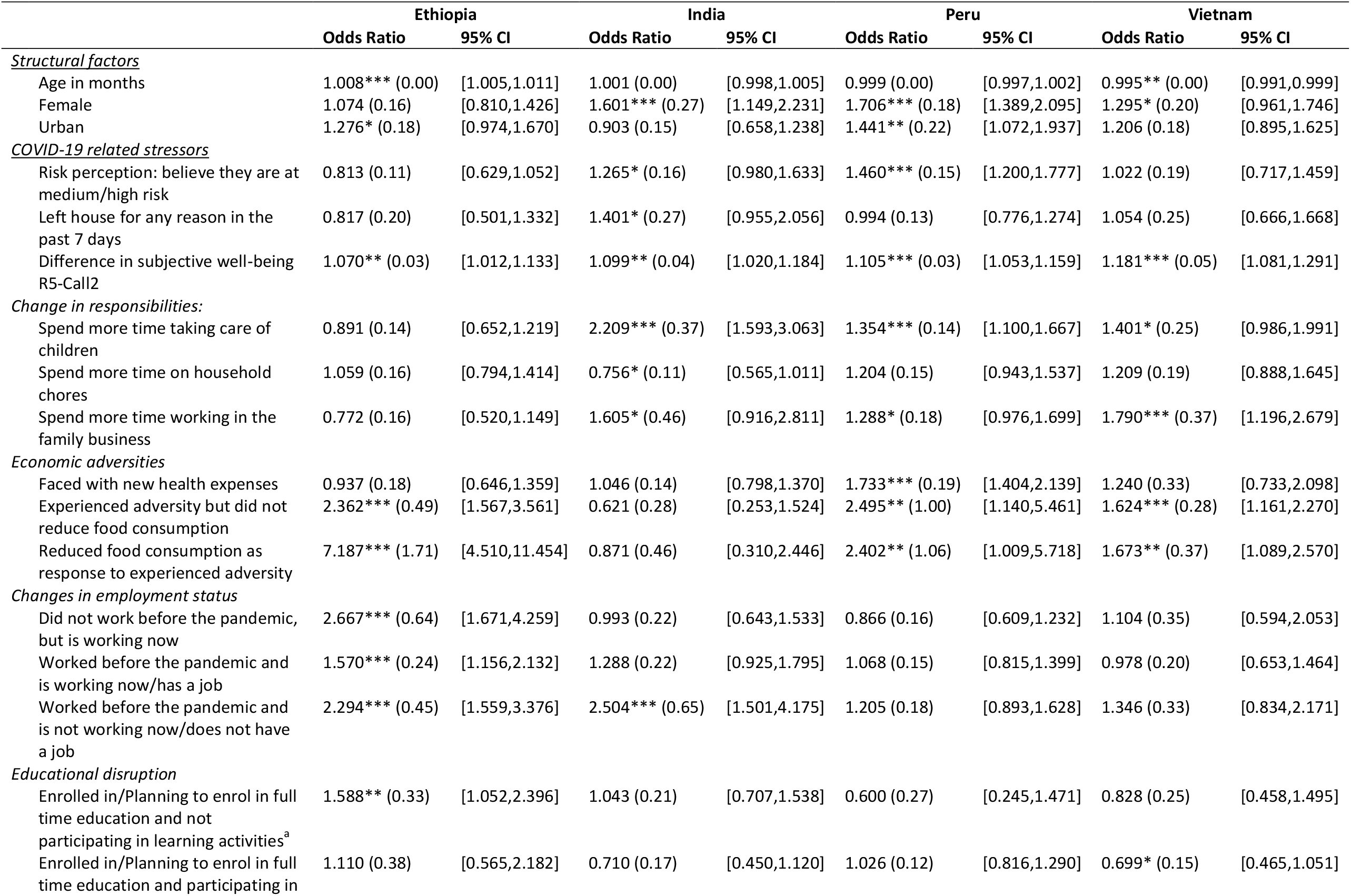
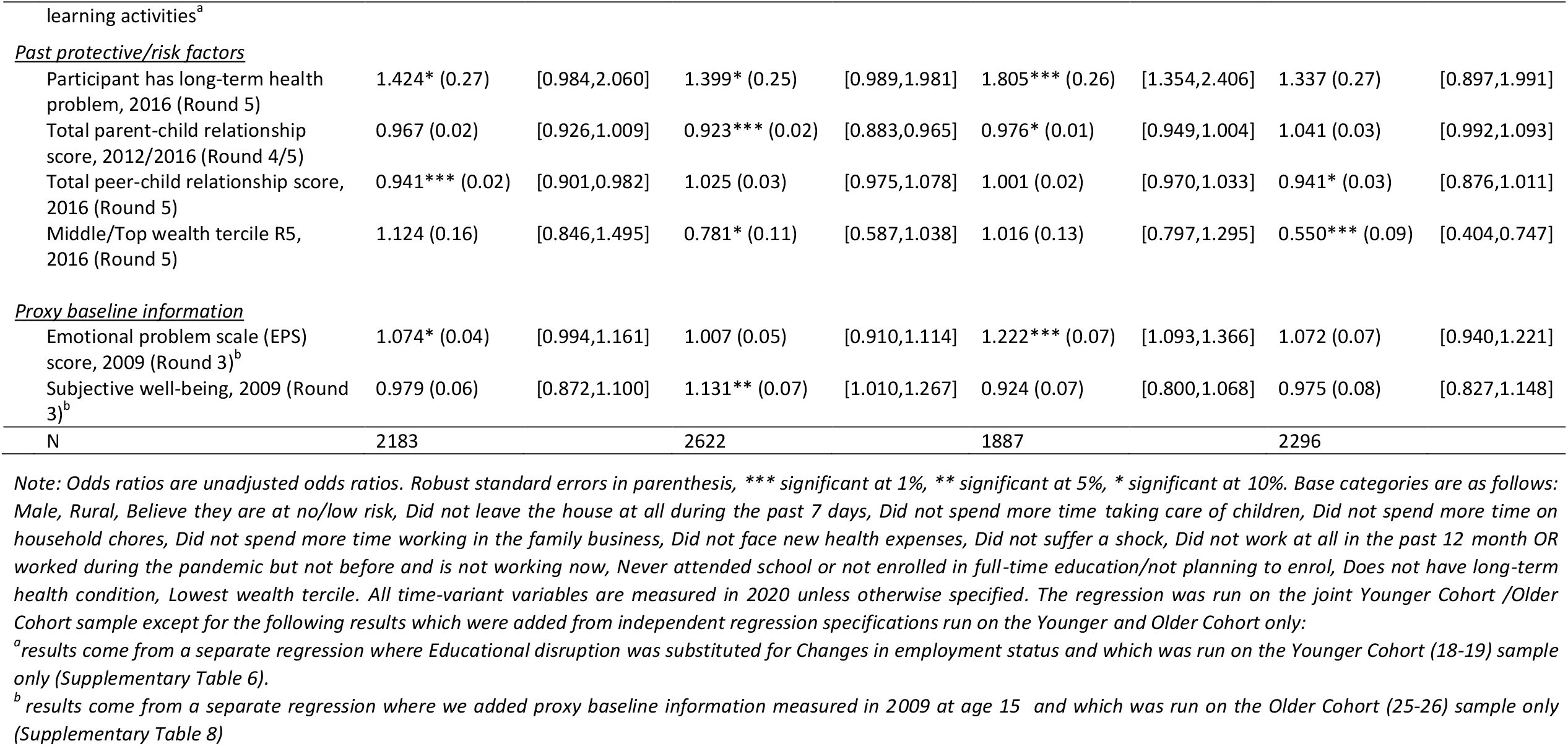
Logistic regression results: Symptoms of at least mild anxiety.

**Table 3.**
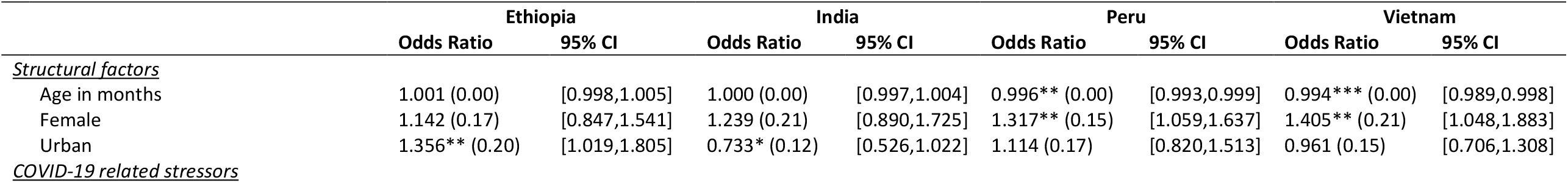
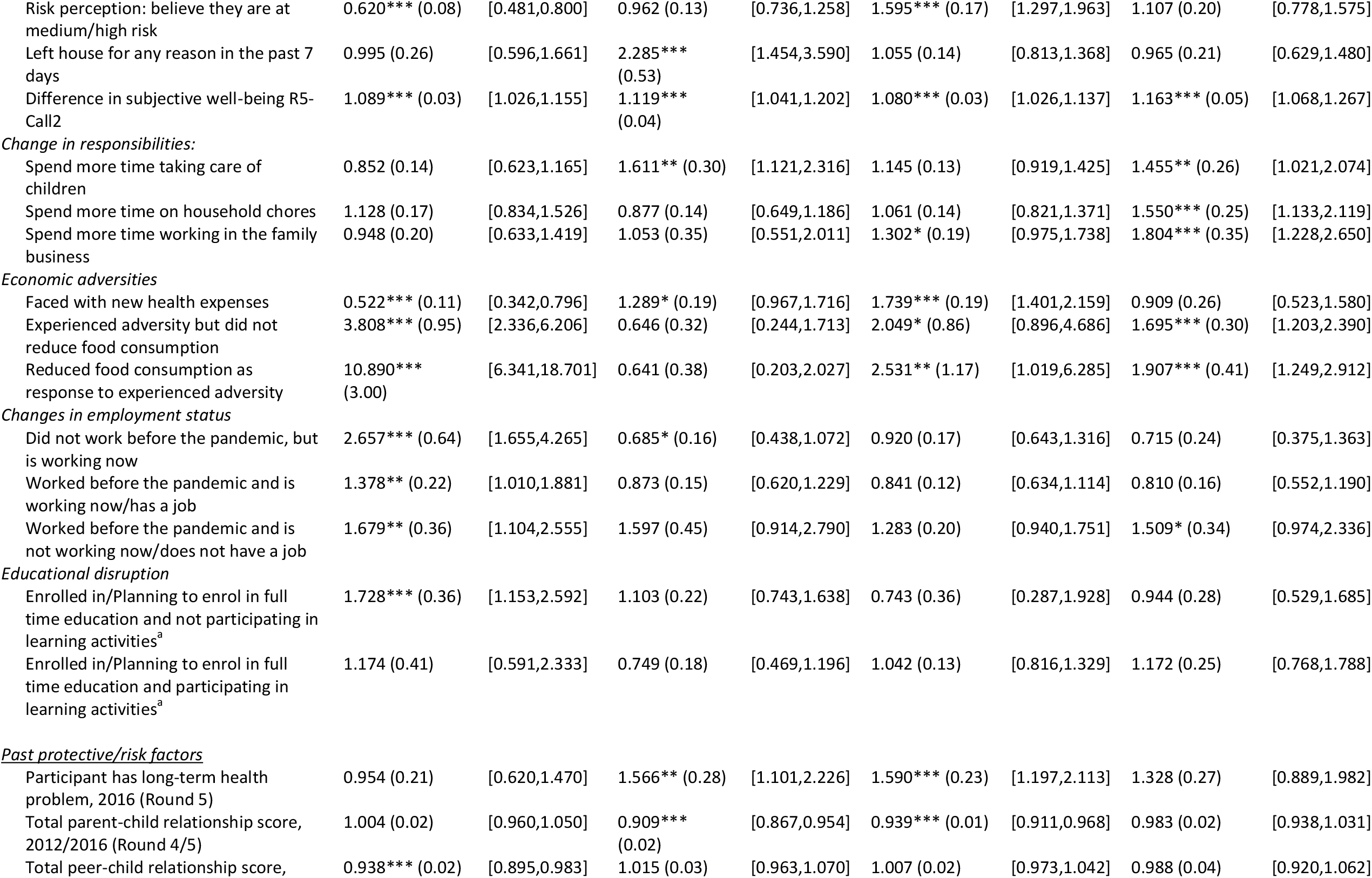
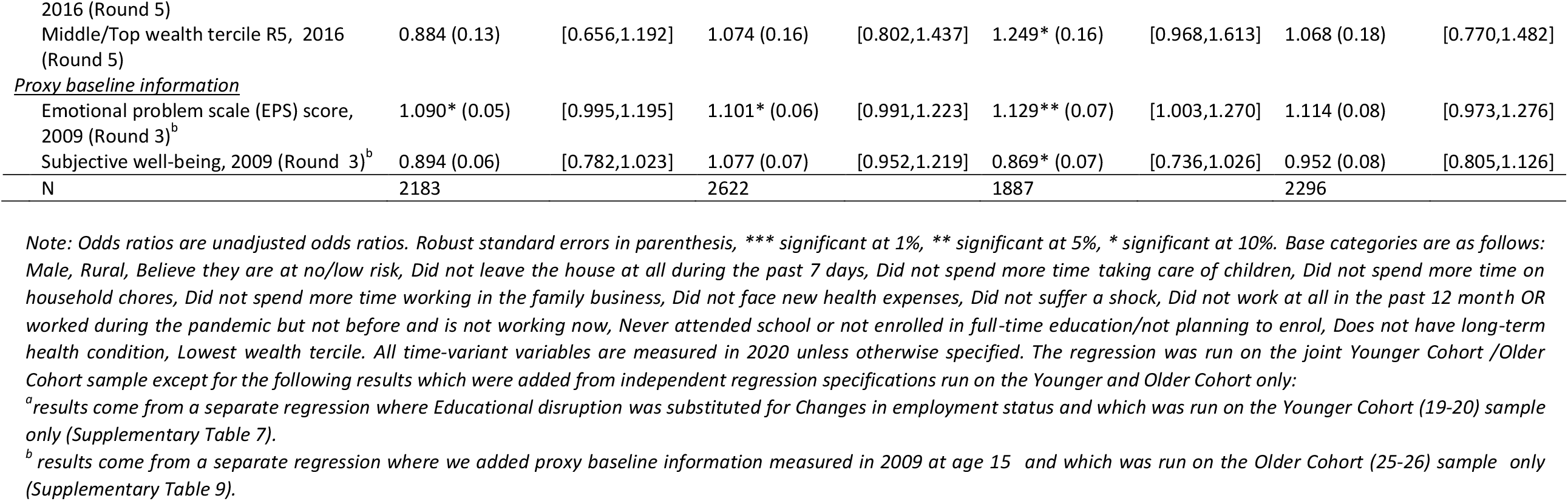
Logistic regression results: Symptoms of at least mild depression.

We noted a high correlation between GAD-7 and PHQ-8 scores (minimum 0·610 (p <0·01) (India) and maximum 0·700 (p <0·01) (Peru), and the rate of having both (at least mild) anxiety and depression symptoms was quite high, with values of up to 24·8% (95% CI 22·87-26·81) in Peru (Supplementary Table 32 and 33).

The significant risk and protective factors were similar. For brevity, the main results refer to associations with experiencing at least mild anxiety symptoms (see **Fehler! Verweisquelle konnte nicht gefunden werden**.) and at the end we comment on differences with results on depression (see **Fehler! Verweisquelle konnte nicht gefunden werden**.) for the sample pooling the two cohorts and females and males together, unless differently specified.

### Structural factors

for females, the odds of at least mild anxiety were 1·30 (95% CI 0·96-1·75, p<0·1) (Vietnam), 1·60 (1·15-2·23, p<0·01) (India) and 1·70 (1·39-2·10, p<0·01) (Peru) times greater than the odds for males (n.s. in Ethiopia). Urban location increased odds significantly in Ethiopia and Peru. Age was not significant in India and Peru, protective in Vietnam, and a risk factor in Ethiopia.

### COVID-related stressors

#### COVID-19 infection risk perception

The odds of those who believed that they were at medium/high risk of catching the virus were 1·27 (95% CI 0·98 - 1·63, p<0·1) (India) to 1·46 (1·20 - 1·78, p<0·01) (Peru) times higher than for those who believed themselves to have no/low risk. The former group had rates of at least mild anxiety of 12% (India) and 45% (Peru).

#### Leaving the house for at least one day a week

No significant effects, except in India where it increased the odds of anxiety by 1·40 (95% CI, 0·96-2·06, p<0·1).

#### Economic adversity

For those who suffered from economic adversity (e.g. fewer clients in the family business, food price increase) odds of anxiety were higher (p<0·01 (Ethiopia and Vietnam), (2·50, 95% CI 1·14-5·46, p<0·05) (2·40, 1·01-5·72, p<0·05) (Peru)) even if it did not cause reduced food consumption. Moreover, in Ethiopia and Vietnam, those who reduced food consumption as a coping strategy had 7·19 (95% CI 4·51-11·45, p<0·01) (Ethiopia), 1·67 (1·09-2·57, p<0·05) (Vietnam) higher odds than those who experienced an adverse event but did not need to reduce food consumption in response 2·36 (1·57-3·56, p<0·01) (Ethiopia) and 1·62 (1·16 - 2·27, p<0·01) (Vietnam) (both compared to those who did not experience any adverse event at all). In Ethiopia, 36% of those who reduced food consumption reported at least mild anxiety compared to 7% for those who did not experience an adverse event (p<0·0001). In Ethiopia, odds were higher among females than males, but in Peru and Vietnam significant for males only. Facing new health expenses significantly increased the odds of displaying at least mild symptoms of anxiety by 1·73 (95% CI 1·40 - 2·14) (p <0·01) in Peru. Over half (52%, p-<0.0001) of those who faced new health expenses report at least mild anxiety (although not significant in India or Ethiopia, significant risk factor for females in Vietnam (1·76, 95% CI 0·93-3·31, p<0·1)).

#### Increased responsibilities

Spending more time on childcare during the lockdown increased odds of anxiety by 2·21 (95% CI 1·59-3·06, p<0·01) in India, 1·35 (1·10-1·67, p<0·01) in Peru and 1·4 (0·99-1·99, p<0·1) in Vietnam. Rates of at least mild anxiety for those who spent more time taking care of children were 20% vs 9% (India), 49% vs 37% (Peru) and 13% vs 8% (Vietnam), (all p<0·001). Spending more time on household chores *lowered* odds in India for anxiety (females only). For those who spend more time working in the family business the odds of anxiety were 1·61 (95% CI 0·92-2·81, p<0·1) times higher in India, 1·29 (0·98-1·67, p<0·1, n.s. for females) in Peru and 1·80 (1·20-2·68, p<0·01) in Vietnam (higher odds among males). In Vietnam, those who spent more time working in the family business reported rates of at least mild anxiety at 16% (p<0·0001), the highest among the Vietnamese sample.

#### Change in subjective well-being (SWB)

the large decrease in SWB between 2016 and the pandemic (registered in all countries but Vietnam) is correlated with GAD-7 and PHQ-8 mean scores in the expected direction (Supplementary Table 34). Furthermore, across countries those with at least mild anxiety (depression) reported lower SWB (2016 and 2020) than their counterparts. Moreover, those with at least mild anxiety (depression) reported larger drops in SWB than their non-anxious (non-depressed) counterparts (Supplementary Table 35 and Supplementary Table 36). For each one-point decrease in SWB between 2016 and the pandemic, the odds of at least mild anxiety increased by a factor of 1·07 (95% CI 1·01-1·13, p<0·05) (Ethiopia), 1·10 (1·02-1·18, p<0·05) (India), 1·11 (1·05-1·16, p<0·01) (Peru) and 1·18 (1·08-1·29, p<0·01) (Vietnam).

#### Changes in employment status

In Ethiopia, those who participated in the labour market had higher odds of anxiety than those who did not (e.g. full-time students, stay-at-home parents). However, the odds of those who were pushed into the labour market (2·67, 95% CI 1·67-4·26, p<0·01) or lost their job (2·29, 1·56-3·38, p<0·01) were higher than those who simply participated (1·57, 1·16-2·13, p<0·01) (all in comparison to non-participants). In India losing a job increased risk of anxiety by 2·50 (95% CI 1·50-4·18, p<0·01). In Peru and Vietnam, there were no employment effects. Rates of at least mild anxiety among those who lost their jobs were among the highest in each country 31% (Ethiopia, p<0·001), 20% (India, p<0·001), 46% (Peru, n.s.), 12% (Vietnam, n.s.).

#### Educational disruption (19-year-old cohort only)

Students who were enrolled in Ethiopia before the pandemic and were unable to access virtual classes, or complete homework had 1·59 (95% CI 1·05-2·40, p<0·05) times higher odds of anxiety than those who were not enrolled. In Vietnam, those who were enrolled and engaged in learning activities had lower odds of anxiety (0·70, 95% CI 0·47-1·05, p<0·1) than those who were not enrolled (base category). The full YC only regression results can be found in the Supplementary Tables 6 and 7; the education results split by gender (again YC only) are located in the Supplementary Tables 20-23.

### Past protective/risk factors

#### Long-term health problems (measured in 2016)

The odds of at least mild anxiety were 1·42 (95% CI 0·98-2·06, p<0·1) (Ethiopia), 1·40 (1·00-2·00, p<0·1) (India) and 1·80 (1·35-2·41, p<0·01) (Peru) times as large as the odds for those who did not (n.s. in Vietnam). In Peru, those reporting long-term health problems had the highest rates of at least mild anxiety, 56% (p<0·0001).

*Parent-child relationship* (measured at ages 15 for the younger cohort, in 2016 and measured at age 19 for the older cohort, in 2013) *and peer-child relationship* (at ages 15 for the younger cohort and at age 22 for the older cohort, in 2016): Strong parent-child relationships were a significant protective factor in India and Peru, while peer-child relationships were a significant protective factor in Ethiopia and Vietnam.

#### Past household wealth (measured in 2016)

i.e. being in the middle/highest wealth tercile vs the lowest was a marginally significant protective factor in India (0·78, 95% CI 0·59-1·04, p<0·1) and significant in Vietnam (0·55, 0·40-0·75, p<0·01).

### Proxy baseline information

*Past emotional problems and well-being* (25-year-old cohort only): For a one-point increase in previous emotional problems at age 15 (measured in 2009), the odds of at least mild anxiety increased by a factor of 1·22 (1·09 - 1·37, p <0·01) (Peru) and 1·07 (95% CI 0·99 - 1·16, p <0·1) (Ethiopia). Notably, the effect of the COVID-19 related stressors holds when controlling for past proxy baseline information. The full older cohort only regression results can be found in Supplementary Tables 8 and 9.

### Significant differences between anxiety and depression regression results

(See ***Fehler! Verweisquelle konnte nicht gefunden werden***.). In Ethiopia, food insecurity had a higher impact on depression for males than females. In India, subjective high infection risk increased anxiety, but not depression, and those faced with new health expenses had higher odds of depression, but not anxiety. Females had higher rates of anxiety but not depression. In Peru, childcare was not a risk factor for depression, and past SWB was a protective factor. In Vietnam losing a job was a significant risk factor for depression, while good peer relations and education were not significant for depression.

## DISCUSSION

We examined the impact of the COVID-19 pandemic on the mental health of young people in Ethiopia, India, Peru and Vietnam. The sample has broad coverage of the poorer population in each country, and we interviewed 93% of those located during the tracking prior the pandemic, including those without internet access and in Ethiopia also those without mobile phone, who would be excluded from an online survey. Internet access has had both positive and negative effects in the pandemic^50^. In our sample, those without access to internet have significantly higher rates of anxiety in Vietnam and Peru.

The four countries have had different experiences of the pandemic – Peru is the most affected country in terms of deaths per population, and Vietnam the least. India, Peru and Vietnam imposed strict lockdowns to control the spread of the virus, and Ethiopia restricted certain activities such as school closure but did not impose a strict lockdown, though faced other challenges (locust infestations, food price inflation and violence).

This study reveals a strong relationship between the severity of the pandemic and the rates of mental health conditions in our sample, both in terms of anxiety and depression symptoms. Rates of at least mild anxiety (depression) were four (three) times higher in Peru compared to Vietnam. Furthermore, the 2020 survey showed a significant fall in subjective wellbeing from 2016 in all countries except Vietnam. The fall in subjective wellbeing is highly correlated with anxiety and depression symptoms. A strong correlation between SWB and our mental health indicators is important as in absence of baseline measures of GAD-7 and PhQ-8, it suggests that SWB is a useful proxy baseline.

The economic impact of the pandemic has affected certain groups of young people in all study countries, even Vietnam, and Ethiopia where there was no full national lockdown. Overall, our findings confirmed that those experiencing COVID-19 related stressors had worse mental health, although the relative importance of stressors varied across countries: increased health expenses and believing they were at a medium/high infection risk was detrimental for young people in Peru, but increased food insecurity was much more important in Ethiopia, reflecting high rates of food price inflation in 2019 which continued into 2020. Moreover, peer relations in earlier years was a protective factor for anxiety and depression only in Ethiopia. In Peru and Vietnam there were no employment effects on anxiety, likely for very different reasons – in Peru health concerns were more important, and in Vietnam the labour market was relatively resilient.

Exploiting the longitudinal data allowed us to investigate past protective/risk factors. As expected, parent and peer relations measured during childhood and adolescence were protective, though in different ways across countries. Strong parental relationships were a significant protective factor in India and Peru, whereas peer relationships were more important in Ethiopia and Vietnam. Those reporting long term health problems were twice as likely to display symptoms consistent with at least mild anxiety, this effect being particularly pronounced in Peru. Previous relative wealth was a significant protective factor only in India and Vietnam. Pre-pandemic emotional problems were risk factors, especially in Ethiopia and Peru. The associations with COVID-19 related stressors were robust to the inclusion of pre-pandemic emotional problems and past subjective well-being.

Other studies used longitudinal data to document the impact of the pandemic on mental health,^5 51^ though none investigated a general population of young people of a similar age in our study countries that we can directly compare to, though results from UK studies have similar findings. The closest study to ours, a phone survey in a developing country finds a deterioration in maternal mental health in rural Bangladesh.^51^ Our study shows lower rates of anxiety and depression in rural areas in Ethiopia and Peru, but significantly higher in India. We are able to disaggregate the effect of a range of COVID-19 related stressors, which we can relate individually to other studies. A study in Hubei province, China ^11^ showed the importance of income losses during the pandemic. Studies of college students in China^8^ and Bangladesh^10^ show that educational disruption significantly increased anxiety and depression, similar to our results in Vietnam and Ethiopia. Social support was negatively correlated with the level of anxiety^8^, similar to our findings regarding parent/peer-child relationships. In Jordan^9^ female healthcare professionals, female university students and university students with chronic disease were at higher risk of developing depression, similar to our results for long-term health problems.

Even controlling for other factors, we found females to be more vulnerable to anxiety in India, Peru and Vietnam and more vulnerable to depression in Peru and Vietnam. In Ethiopia we found no significant gender effects. While most studies^5 9 11^ have similar findings regarding females, one^8^ finds no gender differences among Chinese college students while male Bangladeshi students^10^ had higher depressive symptoms than females.

### Strengths and limitations

Other studies have used longitudinal data to document the impact of the pandemic on mental health.^5, 12^ This study’s strength combines survey data about experiences of COVID-19 with long term information from two cohorts of participants of a population-based cohort study in four LMICs. The study was able to cover the poorest, those without internet (or mobile phone), and examine a broad range of pandemic-related stressors as well as controlling for longer-term risk and protective factors. Our study has a number of limitations. We do not have a directly comparable pre-COVID baseline for depression/anxiety. However, we use proxy variables for baseline and our explanatory variables capture dynamics during the pandemic. Finally, as for other studies, there may be underreporting because of stigma associated with mental health, despite piloting and validation; however, note that our analysis identifies high risk groups *within* each country. Also self-reported variables may be biased due to feelings of anxiety or depression. The findings are not fully generalizable to the whole population of LMICs due to the pro poor design and age group, however they broadly represent poor young people in the study countries.

## CONCLUSION

Adolescents and young people have been lower priority for COVID-19 interventions, given the lower rates of hospitalization and death for this age group. This research shows that the pandemic is having important effects on the mental health of certain groups of young people even in countries with fewer cases. Mental health services are very limited in LMICs making it urgent to develop evidence-based and sustainable prevention programmes in response to the pandemic.

## Supporting information

Research checklist following STROBE guidelines

## Data Availability

The entire individual participant data collected during the phone survey and previous in-person rounds, after de-identification, is available including data dictionaries. Furthermore the questionnaire, attrition reports and the field work manual are available at https://www.younglives.org.uk/. The data will be available in January 2021, with no end date to anyone who wishes to access the data for any purpose, via the UK Data Archive (study number 8678, DOI: 10.5255/UKDA-SN-8678-1).

http://doi.org/10.5255/UKDA-SN-8678-2

## Acknowledgments

We particularly wish to thank the Young Lives respondents and their families for generously giving us their time and cooperation during this difficult time as well as fieldworkers and field managers in the four study countries. We are gratefully indebted to Professor George Patton for his helpful comments.

## Author Contributions

CP and MF conceived the study. CP, MF, DS, and ASJ designed the study. MF, ASJ, RE, TW, LTD led data collection. CP and AH did the statistical analyses. CP, AH and MF wrote the first draft of the Article. AS and MK provided comments and input to the several drafts of the article. MF, DS, ASJ verified the underlying data. AS, MF, CP and AH assessed scale validation and methodology. AH prepared the supplementary files. All authors critically reviewed multiple versions of the manuscript and approved the final version. The authors alone are responsible for the views expressed in this article.

## Patient consent for publication

Not required.

