## Supplemental Material for "Impact of the COVID-19 Pandemic on anxiety and depression symptoms of young people in the Global South: evidence from a four-country cohort study"

### Supplementary Table 1. Sample size

| | Ethiopia | | | India | | | Peru | | | Vietnam | | | Total | | |
|---|---|---|---|---|---|---|---|---|---|---|---|---|---|---|---|
| | Full sample | YC | OC | Full sample | YC | OC | Full sample | YC | OC | Full sample | YC | OC | Full sample | YC | OC |
| Round 5, 2016 | 2626 | 1812 | 814 | 2822 | 1900 | 922 | 2468 | 1860 | 608 | 2848 | 1938 | 910 | 10764 | 7510 | 3254 |
| Tracking samples, 2019-2020 | 2634 | 1803 | 831 | 2815 | 1902 | 913 | 2215 | 1701 | 514 | 2832 | 1922 | 910 | 10496 | 7328 | 3168 |
| Phone survey, 2020 | 2439 | 1665 | 774 | 2754 | 1868 | 886 | 2018 | 1561 | 457 | 2519 | 1691 | 828 | 9730 | 6785 | 2945 |
| Mental health measures available | 2433 | 1659 | 774 | 2753 | 1867 | 886 | 1977 | 1535 | 442 | 2517 | 1690 | 827 | 9680 | 6751 | 2929 |
| Included in our analysis | 2183 | 1533 | 650 | 2622 | 1786 | 836 | 1887 | 1496 | 391 | 2296 | 1596 | 700 | 8988 | 6411 | 2577 |

*Notes: YC stands for Younger Cohort, OC for Older Cohort.*

### Supplementary Table 2. Attrition rates (%)

| | Ethiopia | | | India | | | Peru | | | Vietnam | | | Total | | |
|---|---|---|---|---|---|---|---|---|---|---|---|---|---|---|---|
| | Full sample | YC | OC | Full sample | YC | OC | Full sample | YC | OC | Full sample | YC | OC | Full sample | YC | OC |
| Tracking sample to phone survey | 7.4 | 7.7 | 6.9 | 2.2 | 1.8 | 3.0 | 8.9 | 8.2 | 11.1 | 11.1 | 12.0 | 9.0 | 7.3 | 7.4 | 7.0 |

### Supplementary Table 3. Non-response rate to mental health questions (%)

| Ethiopia | | | India | | | Peru | | | Vietnam | | | Total | | |
|---|---|---|---|---|---|---|---|---|---|---|---|---|---|---|
| Full sample | YC | OC | Full sample | YC | OC | Full sample | YC | OC | Full sample | YC | OC | Full sample | YC | OC |
| 0.2 | 0.4 | 0.0 | 0.0 | 0.1 | 0.0 | 2.0 | 1.7 | 3.3 | 0.1 | 0.1 | 0.1 | 0.5 | 0.5 | 0.5 |

### Supplementary Table 4. Measures of reliability and internal consistency

| | | Cronbach's Alpha | Inter-item correlation | Kaiser–Meyer–Olkin measure of sampling adequacy |
|---|---|---|---|---|
| **Ethiopia** | GAD-7 | 0.728 | 0.277 | 0.792 |
| | PHQ-8 | 0.652 | 0.190 | 0.744 |
| **India** | GAD-7 | 0.691 | 0.242 | 0.777 |
| | PHQ-8 | 0.637 | 0.180 | 0.744 |
| **Peru** | GAD-7 | 0.804 | 0.369 | 0.872 |
| | PHQ-8 | 0.773 | 0.299 | 0.839 |
| **Vietnam** | GAD-7 | 0.749 | 0.299 | 0.814 |
| | PHQ-8 | 0.748 | 0.271 | 0.807 |

*Note: Results are for the combined Younger Cohort/ Older Cohort sample.*

### Supplementary Table 5. Sample description

| | Ethiopia | | India | | Peru | | Vietnam | |
|---|---|---|---|---|---|---|---|---|
| | Mean | sd | Mean | sd | Mean | sd | Mean | sd |
| *Structural factors* | | | | | | | | |
| Female | 46.27 | 0.499 | 47.79 | 0.500 | 49.28 | 0.500 | 51.61 | 0.500 |
| Urban | 45.99 | 0.499 | 28.60 | 0.452 | 81.13 | 0.391 | 45.73 | 0.498 |
| *COVID-19 related stressors* | | | | | | | | |
| Risk perception: believe they are at medium/high risk | 69.31 | 0.461 | 46.38 | 0.499 | 54.27 | 0.498 | 21.39 | 0.410 |
| Left house for any reason in the past 7 days | 94.50 | 0.228 | 79.25 | 0.406 | 80.45 | 0.397 | 87.67 | 0.329 |
| Difference in subjective well-being 2016-2020 | 0.96 | 2.068 | 0.49 | 1.949 | 0.52 | 2.066 | -0.39 | 1.876 |
| *Change in responsibilities:* | | | | | | | | |
| Spend more time taking care of children | 29.00 | 0.454 | 18.31 | 0.387 | 34.23 | 0.475 | 22.91 | 0.420 |
| Spend more time on household chores | 46.82 | 0.499 | 50.95 | 0.500 | 77.53 | 0.417 | 51.44 | 0.500 |
| Spend more time working in the family business | 12.96 | 0.336 | 4.12 | 0.199 | 15.00 | 0.357 | 10.89 | 0.312 |
| *Economic adversities* | | | | | | | | |
| Faced with new health expenses | 10.12 | 0.302 | 55.95 | 0.497 | 29.57 | 0.456 | 6.53 | |
| Did not experience adversity | 20.71 | 0.405 | 1.72 | 0.130 | 2.60 | 0.159 | 42.25 | 0.494 |
| Experienced adversity but did not reduce food consumption | 65.83 | 0.474 | 92.68 | 0.261 | 90.94 | 0.287 | 41.94 | 0.494 |
| Reduced food consumption as response to experienced adversity | 13.47 | 0.341 | 5.61 | 0.230 | 6.47 | 0.246 | 15.81 | 0.365 |
| *Changes in employment status* | | | | | | | | |
| Did not work at all in the past 12 month OR worked during the pandemic but not before the pandemic (and is not working now) | 46.17 | 0.499 | 41.38 | 0.493 | 24.06 | 0.428 | 23.48 | 0.424 |
| Did not work before the pandemic, but is working now | 6.83 | 0.252 | 16.40 | 0.370 | 12.35 | 0.329 | 6.75 | 0.251 |
| Worked before the pandemic and is working now/has a job | 38.71 | 0.487 | 37.57 | 0.484 | 46.74 | 0.499 | 56.36 | 0.496 |
| Worked before the pandemic and is not working now/does not have a job | 8.29 | 0.276 | 4.65 | 0.211 | 16.85 | 0.374 | 13.41 | 0.341 |
| *Educational disruption* | | | | | | | | |
| Never attended school or not enrolled in full-time education/not planning to enrol[a] | 32.68 | 0.469 | 33.87 | 0.473 | 58.96 | 0.492 | 47.49 | 0.500 |
| Enrolled in/Planning to enrol in full time education and not participating in learning activities[a] | 58.71 | 0.493 | 36.28 | 0.481 | 1.87 | 0.136 | 12.72 | 0.333 |
| Enrolled in/Planning to enrol in full time education and participating in learning activities[a] | 8.61 | 0.281 | 29.84 | 0.458 | 39.17 | 0.488 | 39.79 | 0.490 |

| | Ethiopia | | India | | Peru | | Vietnam | |
|---|---|---|---|---|---|---|---|---|
| | Mean | sd | Mean | sd | Mean | sd | Mean | sd |
| *Past protective/risk factors* | | | | | | | | |
| Child has long-term health problem, 2016 (Round 5) | 8.80 | 0.283 | 11.48 | 0.319 | 12.67 | 0.333 | 14.20 | 0.349 |
| Total parent-child relationship score, 2012/2016 (Round 4/5) | 26.26 | 2.848 | 27.63 | 2.916 | 24.86 | 3.610 | 25.74 | 3.204 |
| Total peer-child relationship score, 2016 (Round 5) | 24.13 | 2.866 | 25.02 | 2.631 | 23.41 | 3.204 | 22.57 | |
| Middle/Top wealth tercile R5, 2016 (Round 5) | 67.11 | 0.470 | 66.44 | 0.472 | 69.00 | 0.463 | 66.03 | 2.257 |
| *Proxy baseline information* | | | | | | | | |
| Emotional problem scale (EPS) score, 2009 (Round 3)[b] | 2.78 | 2.473 | 3.57 | 2.348 | 4.27 | 2.323 | 3.68 | 2.143 |
| Subjective well-being, 2009 (Round 3)[b] | 4.86 | 1.719 | 4.73 | 1.815 | 6.12 | 1.616 | 5.43 | 1.604 |
| N | 2183 | | 2622 | | 1887 | | 2296 | |

*Note: Mean values are reported. Self-reported risk belief coded the question "chance of getting infected with Coronavirus" into no/low risk, or medium/high risk. All time-variant variables are measured in 2020 unless otherwise specified. [a] refers to Younger Cohort (18-19) only, [b] refers to Older Cohort (25-26) only. Around 30% of the sample in Ethiopia, India and Vietnam are part of the Older Cohort. In Peru they make up 20% of the sample. The average age of the Younger Cohort in Ethiopia, India, Peru, Vietnam in years (months) is 18 (226), 18 (227), 18 (227), and 18 (227). The average age of the Older Cohort in Ethiopia, India, Peru, Vietnam in years (months) is 25 (309), 25 (310), 25 (311), and 25 (311).*

**Supplementary Table 6. Logistic regression results: Symptoms of at least mild anxiety (Younger Cohort only)**

| | Ethiopia | | India | | Peru | | Vietnam | |
|---|---|---|---|---|---|---|---|---|
| | **Odds Ratio** | **95% CI** | **Odds Ratio** | **95% CI** | **Odds Ratio** | **95% CI** | **Odds Ratio** | **95% CI** |
| *Structural factors* | | | | | | | | |
| Age in months | 1.000 (0.02) | [0.960,1.042] | 0.987 (0.02) | [0.946,1.030] | 0.991 (0.01) | [0.963,1.020] | 1.020 (0.03) | [0.967,1.077] |
| Female | 1.216 (0.21) | [0.874,1.692] | 1.420* (0.30) | [0.940,2.143] | 1.758*** (0.20) | [1.402,2.205] | 1.453** (0.25) | [1.034,2.041] |
| Urban | 1.565** (0.31) | [1.059,2.312] | 1.198 (0.24) | [0.806,1.781] | 1.362* (0.22) | [0.986,1.882] | 1.322 (0.26) | [0.901,1.940] |
| *COVID-19 related changes/behaviours* | | | | | | | | |
| Risk perception: believe they are at medium/high risk | 0.679** (0.12) | [0.487,0.947] | 1.244 (0.21) | [0.898,1.724] | 1.432*** (0.16) | [1.149,1.784] | 1.031 (0.22) | [0.673,1.579] |
| Left house for any reason in the past 7 days | 0.885 (0.27) | [0.482,1.627] | 1.576* (0.41) | [0.948,2.620] | 1.023 (0.14) | [0.784,1.335] | 1.304 (0.34) | [0.783,2.171] |
| Difference in subjective well-being between Round 5 (2016) and phone survey (2020) | 1.043 (0.04) | [0.969,1.123] | 1.103** (0.05) | [1.007,1.207] | 1.102*** (0.03) | [1.046,1.161] | 1.195*** (0.06) | [1.084,1.316] |
| *Change in responsibilities:* | | | | | | | | |
| Spend more time taking care of children | 1.012 (0.20) | [0.692,1.479] | 2.176*** (0.52) | [1.363,3.475] | 1.509*** (0.18) | [1.194,1.906] | 1.400 (0.31) | [0.913,2.147] |
| Spend more time on household chores | 0.983 (0.17) | [0.694,1.392] | 0.816 (0.15) | [0.570,1.169] | 1.180 (0.17) | [0.895,1.557] | 1.013 (0.18) | [0.718,1.430] |
| Spend more time working in the family business | 0.703 (0.21) | [0.394,1.253] | 2.117** (0.67) | [1.136,3.943] | 1.235 (0.19) | [0.917,1.663] | 1.640** (0.38) | [1.037,2.593] |
| *Economic shocks* | | | | | | | | |
| Faced with new health expenses | 0.668 (0.21) | [0.360,1.240] | 0.881 (0.16) | [0.624,1.245] | 1.741*** (0.21) | [1.371,2.209] | 1.309 (0.42) | [0.699,2.455] |
| Experienced adversity but did not reduce food consumption | 3.636*** (0.99) | [2.128,6.213] | 0.702 (0.38) | [0.245,2.011] | 2.503** (1.08) | [1.074,5.834] | 1.493** (0.29) | [1.021,2.184] |
| Reduced food consumption as response to experienced adversity | 8.158*** (2.58) | [4.393,15.149] | 1.173 (0.73) | [0.345,3.985] | 2.154 (1.03) | [0.844,5.498] | 1.680** (0.42) | [1.027,2.748] |
| *Educational disruption* | | | | | | | | |
| Enrolled in/Planning to enrol in full time education and not participating in learning activities | 1.588** (0.33) | [1.052,2.396] | 1.043 (0.21) | [0.707,1.538] | 0.600 (0.27) | [0.245,1.471] | 0.828 (0.25) | [0.458,1.495] |
| Enrolled in/Planning to enrol in full time education and participating in learning activities | 1.110 (0.38) | [0.565,2.182] | 0.710 (0.17) | [0.450,1.120] | 1.026 (0.12) | [0.816,1.290] | 0.699* (0.15) | [0.465,1.051] |
| *Past protective/risk factors* | | | | | | | | |
| Participant has long-term health problem, 2016 (Round 5) | 1.298 (0.36) | [0.755,2.233] | 1.226 (0.30) | [0.754,1.995] | 1.624*** (0.28) | [1.163,2.266] | 1.030 (0.29) | [0.591,1.793] |

| | Ethiopia | | India | | Peru | | Vietnam | |
|---|---|---|---|---|---|---|---|---|
| | Odds Ratio | 95% CI | Odds Ratio | 95% CI | Odds Ratio | 95% CI | Odds Ratio | 95% CI |
| Total parent-child relationship score, 2012/2016 (Round 4/5) | 0.959 (0.03) | [0.902,1.019] | 0.912*** (0.03) | [0.861,0.967] | 0.975 (0.02) | [0.944,1.007] | 1.030 (0.03) | [0.976,1.088] |
| Total peer-child relationship score, 2016 (Round 5) | 0.951 (0.03) | [0.895,1.011] | 1.015 (0.04) | [0.948,1.088] | 0.999 (0.02) | [0.964,1.035] | 0.968 (0.04) | [0.896,1.047] |
| Middle/Top wealth tercile R5, 2016 (Round 5) | 0.908 (0.19) | [0.608,1.356] | 0.712* (0.13) | [0.495,1.023] | 1.105 (0.15) | [0.845,1.444] | 0.677** (0.13) | [0.467,0.981] |
| N | 1533 | | 1786 | | 1496 | | 1596 | |

*Note: Odds ratios are unadjusted odds ratios. Robust standard errors in parenthesis, *** significant at 1%, ** significant at 5%, * significant at 10%. Base categories are as follows: Male, Rural, Believe they are at no/low risk, Did not leave the house at all during the past 7 days, Did not spend more time taking care of children, Did not spend more time on household chores, Did not spend more time working in the family business, Did not face new health expenses, Did not suffer a shock, Never attended school or not enrolled in full-time education/not planning to enrol, Does not have long-term health condition, Lowest wealth tercile. All time-variant variables are measured in 2020 unless otherwise specified.*

### Supplementary Table 7. Logistic regression results: Symptoms of at least mild depression (Younger Cohort only)

| | Ethiopia | | India | | Peru | | Vietnam | |
|---|---|---|---|---|---|---|---|---|
| | Odds Ratio | 95% CI | Odds Ratio | 95% CI | Odds Ratio | 95% CI | Odds Ratio | 95% CI |
| *Structural factors* | | | | | | | | |
| Age in months | 0.999 (0.02) | [0.959,1.041] | 0.997 (0.02) | [0.954,1.041] | 0.974* (0.01) | [0.945,1.003] | 1.028 (0.03) | [0.975,1.084] |
| Female | 1.318 (0.23) | [0.940,1.848] | 1.225 (0.25) | [0.821,1.828] | 1.282** (0.16) | [1.009,1.630] | 1.438** (0.24) | [1.032,2.004] |
| Urban | 1.533** (0.29) | [1.051,2.235] | 0.969 (0.20) | [0.644,1.457] | 1.175 (0.20) | [0.840,1.643] | 1.093 (0.22) | [0.736,1.623] |
| *COVID-19 related changes/behaviours* | | | | | | | | |
| Risk perception: believe they are at medium/high risk | 0.498*** (0.08) | [0.361,0.687] | 0.966 (0.16) | [0.693,1.346] | 1.548*** (0.18) | [1.230,1.949] | 1.025 (0.22) | [0.674,1.559] |
| Left house for any reason in the past 7 days | 0.940 (0.28) | [0.520,1.702] | 2.537*** (0.77) | [1.403,4.589] | 0.928 (0.13) | [0.704,1.223] | 1.163 (0.28) | [0.724,1.867] |
| Difference in subjective well-being between Round 5 (2016) and phone survey (2020) | 1.049 (0.04) | [0.973,1.130] | 1.102** (0.05) | [1.011,1.201] | 1.079*** (0.03) | [1.020,1.142] | 1.169*** (0.06) | [1.065,1.284] |
| *Change in responsibilities:* | | | | | | | | |
| Spend more time taking care of children | 1.061 (0.20) | [0.736,1.527] | 2.170*** (0.51) | [1.370,3.438] | 1.268* (0.16) | [0.992,1.621] | 1.616** (0.33) | [1.087,2.403] |
| Spend more time on household chores | 1.082 (0.19) | [0.763,1.533] | 0.857 (0.16) | [0.592,1.241] | 1.142 (0.17) | [0.858,1.520] | 1.840*** (0.34) | [1.287,2.631] |
| Spend more time working in the family business | 1.076 (0.29) | [0.636,1.823] | 0.803 (0.34) | [0.354,1.823] | 1.160 (0.18) | [0.850,1.582] | 1.724** (0.38) | [1.121,2.652] |

| | Ethiopia | | India | | Peru | | Vietnam | |
|---|---|---|---|---|---|---|---|---|
| | **Odds Ratio** | **95% CI** | **Odds Ratio** | **95% CI** | **Odds Ratio** | **95% CI** | **Odds Ratio** | **95% CI** |
| *Economic shocks* | | | | | | | | |
| Faced with new health expenses | 0.222*** (0.10) | [0.093,0.530] | 1.062 (0.19) | [0.747,1.510] | 1.851*** (0.23) | [1.450,2.363] | 0.805 (0.28) | [0.410,1.580] |
| Experienced adversity but did not reduce food consumption | 4.913*** (1.49) | [2.712,8.902] | 0.651 (0.36) | [0.218,1.944] | 2.541* (1.21) | [0.997,6.476] | 1.763*** (0.36) | [1.181,2.631] |
| Reduced food consumption as response to experienced adversity | 12.771*** (4.42) | [6.484,25.156] | 0.913 (0.61) | [0.248,3.357] | 3.620** (1.89) | [1.300,10.079] | 2.292*** (0.57) | [1.410,3.726] |
| *Educational disruption* | | | | | | | | |
| Enrolled in/Planning to enrol in full time education and not participating in learning activities | 1.728*** (0.36) | [1.153,2.592] | 1.103 (0.22) | [0.743,1.638] | 0.743 (0.36) | [0.287,1.928] | 0.944 (0.28) | [0.529,1.685] |
| Enrolled in/Planning to enrol in full time education and participating in learning activities | 1.174 (0.41) | [0.591,2.333] | 0.749 (0.18) | [0.469,1.196] | 1.042 (0.13) | [0.816,1.329] | 1.172 (0.25) | [0.768,1.788] |
| *Past protective/risk factors* | | | | | | | | |
| Participant has long-term health problem, 2016 (Round 5) | 0.540* (0.20) | [0.265,1.098] | 1.461 (0.35) | [0.909,2.346] | 1.502** (0.26) | [1.077,2.095] | 1.466 (0.37) | [0.889,2.415] |
| Total parent-child relationship score, 2012/2016 (Round 4/5) | 1.011 (0.03) | [0.954,1.072] | 0.890*** (0.03) | [0.838,0.945] | 0.924*** (0.02) | [0.892,0.957] | 0.971 (0.03) | [0.918,1.026] |
| Total peer-child relationship score, 2016 (Round 5) | 0.902*** (0.03) | [0.848,0.960] | 1.000 (0.04) | [0.930,1.076] | 1.025 (0.02) | [0.987,1.065] | 0.987 (0.04) | [0.910,1.071] |
| Middle/Top wealth tercile R5, 2016 (Round 5) | 0.763 (0.15) | [0.518,1.122] | 1.275 (0.24) | [0.879,1.851] | 1.166 (0.17) | [0.878,1.550] | 1.189 (0.24) | [0.797,1.773] |
| N | 1533 | | 1786 | | 1496 | | 1596 | |

*Note: Odds ratios are unadjusted odds ratios. Robust standard errors in parenthesis, *** significant at 1%, ** significant at 5%, * significant at 10%. Base categories are as follows: Male, Rural, Believe they are at no/low risk, Did not leave the house at all during the past 7 days, Did not spend more time taking care of children, Did not spend more time on household chores, Did not spend more time working in the family business, Did not face new health expenses, Did not suffer a shock, Never attended school or not enrolled in full-time education/not planning to enrol, Does not have long-term health condition, Lowest wealth tercile. All time-variant variables are measured in 2020 unless otherwise specified.*

### Supplementary Table 8. Logistic regression results: Symptoms of at least mild anxiety (Older Cohort only)

| | Ethiopia | | India | | Peru | | Vietnam | |
|---|---|---|---|---|---|---|---|---|
| | **Odds Ratio** | **95% CI** | **Odds Ratio** | **95% CI** | **Odds Ratio** | **95% CI** | **Odds Ratio** | **95% CI** |
| <u>*Structural factors*</u> | | | | | | | | |
| Age in months | 1.008 (0.03) | [0.956,1.063] | 1.052* (0.03) | [0.999,1.108] | 0.977 (0.03) | [0.923,1.034] | 0.960 (0.04) | [0.879,1.047] |
| Female | 0.754 (0.21) | [0.432,1.319] | 1.954** (0.58) | [1.087,3.511] | 1.125 (0.30) | [0.670,1.888] | 0.790 (0.27) | [0.404,1.545] |
| Urban | 1.087 (0.23) | [0.713,1.658] | 0.561** (0.15) | [0.334,0.942] | 1.825 (0.75) | [0.819,4.065] | 0.961 (0.31) | [0.516,1.790] |
| <u>*COVID-19 related changes/behaviours*</u> | | | | | | | | |
| Risk perception: believe they are at medium/high risk | 1.466 (0.35) | [0.915,2.346] | 1.383 (0.30) | [0.908,2.108] | 1.693** (0.41) | [1.051,2.728] | 1.060 (0.37) | [0.536,2.098] |
| Left house for any reason in the past 7 days | 0.693 (0.33) | [0.274,1.751] | 1.257 (0.38) | [0.697,2.267] | 0.850 (0.31) | [0.414,1.746] | 0.432* 0.21) | [0.169,1.109] |
| Difference in subjective well-being between Round 5 (2016) and phone survey (2020) | 1.089* (0.05) | [0.987,1.201] | 1.121* (0.07) | [0.983,1.278] | 1.132* (0.08) | [0.990,1.294] | 1.105 (0.12) | [0.901,1.356] |
| *Change in responsibilities:* | | | | | | | | |
| Spend more time taking care of children | 0.735 (0.21) | [0.420,1.285] | 2.081***(0.52) | [1.271,3.405] | 0.971 (0.25) | [0.590,1.599] | 1.230 (0.46) | [0.595,2.544] |
| Spend more time on household chores | 1.313 (0.38) | [0.742,2.325] | 0.639* (0.16) | [0.394,1.034] | 1.667 (0.53) | [0.893,3.114] | 2.240**(0.87) | [1.043,4.808] |
| Spend more time working in the family business | 1.275 (0.41) | [0.682,2.383] | 0.498 (0.41) | [0.100,2.478] | 1.599 (0.54) | [0.829,3.085] | 2.126* (0.96) | [0.879,5.139] |
| *Economic shocks* | | | | | | | | |
| Faced with new health expenses | 1.523 (0.45) | [0.854,2.716] | 1.435 (0.34) | [0.901,2.286] | 1.572* (0.38) | [0.984,2.513] | 0.808 (0.41) | [0.302,2.164] |
| Experienced adversity but did not reduce food consumption | 0.908 (0.31) | [0.466,1.767] | 0.560 (0.49) | [0.102,3.068] | 1.312 (1.77) | [0.093,18.595] | 1.988* (0.78) | [0.922,4.284] |
| Reduced food consumption as response to experienced adversity | 4.323***(1.81) | [1.900,9.835] | 0.567 (0.55) | [0.086,3.751] | 2.691 (3.82) | [0.167,43.423] | 1.986 (0.96) | [0.772,5.113] |
| *Changes in employment status* | | | | | | | | |
| Did not work before the pandemic, but is working now | 2.044 (0.97) | [0.809,5.165] | 0.951 (0.47) | [0.358,2.525] | 1.095 (0.62) | [0.363,3.303] | 0.868 (0.78) | [0.149,5.071] |
| Worked before the pandemic and is working now/has a job | 1.566* (0.39) | [0.956,2.567] | 1.306 (0.36) | [0.756,2.255] | 1.401 (0.56) | [0.638,3.075] | 0.591 (0.34) | [0.194,1.796] |
| Worked before the pandemic and is not working | 2.787***(0.85) | [1.534,5.063] | 2.312** (0.95) | [1.037,5.156] | 5.434***(2.55) | [2.167,13.626] | 1.693 (1.03) | [0.513,5.582] |

| | Ethiopia | | India | | Peru | | Vietnam | |
|---|---|---|---|---|---|---|---|---|
| | Odds Ratio | 95% CI | Odds Ratio | 95% CI | Odds Ratio | 95% CI | Odds Ratio | 95% CI |
| now/does not have a job | | | | | | | | |
| *Past protective/risk factors* | | | | | | | | |
| Participant has long-term health problem, 2016 (Round 5) | 1.134 (0.33) | [0.637,2.019] | 1.583* (0.40) | [0.960,2.612] | 2.591***(0.91) | [1.298,5.171] | 1.647 (0.54) | [0.862,3.150] |
| Total parent-child relationship score, 2012/2016 (Round 4/5) | 0.957 (0.03) | [0.899,1.018] | 0.939* (0.03) | [0.875,1.007] | 0.997 (0.03) | [0.940,1.058] | 1.118* (0.07) | [0.989,1.265] |
| Total peer-child relationship score, 2016 (Round 5) | 0.948 (0.03) | [0.887,1.014] | 1.041 (0.04) | [0.963,1.124] | 1.011 (0.04) | [0.938,1.089] | 0.858* (0.08) | [0.720,1.023] |
| Middle/Top wealth tercile R5, 2016 (Round 5) | 1.033 (0.23) | [0.670,1.594] | 0.871 (0.20) | [0.551,1.376] | 0.635 (0.20) | [0.342,1.180] | 0.488** 0.16) | [0.260,0.914] |
| *Proxy baseline information* | | | | | | | | |
| Emotional problem scale (EPS) score, 2009 (Round 3) | 1.074* (0.04) | [0.994,1.161] | 1.007 (0.05) | [0.910,1.114] | 1.222*** 0.07) | [1.093,1.366] | 1.072 (0.07) | [0.940,1.221] |
| Subjective well-being, 2009 (Round 3) | 0.979 (0.06) | [0.872,1.100] | 1.131** (0.07) | [1.010,1.267] | 0.924 (0.07) | [0.800,1.068] | 0.975 (0.08) | [0.827,1.148] |
| N | 650 | | 836 | | 391 | | 700 | |

*Note: Odds ratios are unadjusted odds ratios. Robust standard errors in parenthesis, *** significant at 1%, ** significant at 5%, * significant at 10%. Base categories are as follows: Male, Rural, Believe they are at no/low risk, Did not leave the house at all during the past 7 days, Did not spend more time taking care of children, Did not spend more time on household chores, Did not spend more time working in the family business, Did not face new health expenses, Did not suffer a shock, Did not work at all in the past 12 month OR worked during the pandemic but not before and is not working now, Does not have long-term health condition, Lowest wealth tercile. All time-variant variables are measured in 2020 unless otherwise specified.*

### Supplementary Table 9. Logistic regression results: Symptoms of at least mild depression (Older Cohort only)

| Depression OC | Ethiopia | | India | | Peru | | Vietnam | |
|---|---|---|---|---|---|---|---|---|
| | Odds Ratio | 95% CI | Odds Ratio | 95% CI | Odds Ratio | 95% CI | Odds Ratio | 95% CI |
| *Structural factors* | | | | | | | | |
| Age in months | 1.018 (0.03) | [0.962,1.077] | 1.065**(0.03) | [1.006,1.127] | 0.984 (0.03) | [0.927,1.044] | 0.985 (0.04) | [0.915,1.060] |
| Female | 0.834 (0.30) | [0.413,1.683] | 1.301 (0.41) | [0.705,2.403] | 1.266 (0.35) | [0.734,2.185] | 1.093 (0.37) | [0.567,2.107] |
| Urban | 1.395 (0.36) | [0.846,2.298] | 0.549**(0.16) | [0.313,0.961] | 1.162 (0.50) | [0.496,2.722] | 0.512**(0.17) | [0.266,0.988] |
| *COVID-19 related changes/behaviours* | | | | | | | | |
| Risk perception: believe they are at medium/high risk | 1.595* (0.43) | [0.942,2.703] | 0.956 (0.22) | [0.607,1.505] | 1.610* 0.41) | [0.980,2.646] | 1.672 (0.58) | [0.851,3.287] |

| Depression | OC | Ethiopia | | India | | Peru | | Vietnam | |
|---|---|---|---|---|---|---|---|---|---|
| | | Odds Ratio | 95% CI | Odds Ratio | 95% CI | Odds Ratio | 95% CI | Odds Ratio | 95% CI |
| | Left house for any reason in the past 7 days | 1.216 (0.69) | [0.402,3.672] | 1.780 (0.63) | [0.890,3.559] | 1.778 (0.78) | [0.753,4.195] | 0.287***(0.14) | [0.113,0.731] |
| | Difference subjective well-being between Round 5 (2016) and phone survey (2020) | 1.221***(0.08) | [1.082,1.377] | 1.175***(0.07) | [1.043,1.324] | 1.076 (0.08) | [0.926,1.250] | 1.129 (0.12) | [0.915,1.393] |
| *Change in responsibilities:* | | | | | | | | | |
| | Spend more time taking care of children | 0.517** (0.16) | [0.279,0.959] | 1.135 (0.33) | [0.637,2.023] | 0.707 (0.20) | [0.401,1.247] | 1.252 (0.50) | [0.571,2.746] |
| | Spend more time on household chores | 1.457 (0.54) | [0.708,3.002] | 0.858 (0.24) | [0.499,1.476] | 1.026 (0.33) | [0.543,1.940] | 0.912 (0.33) | [0.448,1.858] |
| | Spend more time working in the family business | 1.215 (0.42) | [0.614,2.407] | 1.258 (0.73) | [0.402,3.938] | 1.534 (0.53) | [0.779,3.022] | 1.888 (0.87) | [0.763,4.673] |
| *Economic shocks* | | | | | | | | | |
| | Faced with new health expenses | 1.278 (0.43) | [0.657,2.487] | 1.821** (0.48) | [1.089,3.045] | 1.226 (0.31) | [0.745,2.017] | 0.681 (0.39) | [0.223,2.079] |
| | Experienced adversity but did not reduce food consumption | 1.566 (0.69) | [0.661,3.713] | 1.027 (1.15) | [0.114,9.237] | 0.463 (0.57) | [0.042,5.082] | 1.513 (0.55) | [0.744,3.074] |
| | Reduced food consumption as response to experienced adversity | 4.926***(2.50) | [1.825,13.296] | 0.520 (0.66) | [0.042,6.360] | 0.363 (0.48) | [0.027,4.887] | 1.005 (0.51) | [0.374,2.699] |
| *Changes in employment status* | | | | | | | | | |
| | Did not work before the pandemic, but is working now[3] | 3.429***(1.59) | [1.382,8.507] | 0.270* (0.20) | [0.061,1.185] | 1.044 (0.69) | [0.285,3.829] | 1.000 (.) | [1.000,1.000] |
| | Worked before the pandemic and is working now/has a job | 1.588 (0.45) | [0.915,2.758] | 0.740 (0.21) | [0.422,1.297] | 0.943 (0.43) | [0.387,2.299] | 0.634 (0.37) | [0.203,1.978] |
| | Worked before the pandemic and is not working now/does not have a job | 2.360** (0.87) | [1.145,4.865] | 1.267 (0.55) | [0.541,2.968] | 1.986 (1.00) | [0.741,5.323] | 1.747 (1.07) | [0.523,5.832] |
| <u>*Past protective/risk factors*</u> | | | | | | | | | |
| | Participant has long-term health problem, 2016 (Round 5) | 1.129 (0.40) | [0.562,2.268] | 1.636* (0.46) | [0.941,2.844] | 2.246**(0.74) | [1.176,4.291] | 1.036 (0.35) | [0.530,2.025] |

| Depression | OC | Ethiopia | | India | | Peru | | Vietnam | |
|---|---|---|---|---|---|---|---|---|---|
| | | Odds Ratio | 95% CI | Odds Ratio | 95% CI | Odds Ratio | 95% CI | Odds Ratio | 95% CI |
| | Total parent-child relationship score, 2012/2016 (Round 4/5) | 1.003 (0.04) | [0.934,1.078] | 0.953 (0.04) | [0.877,1.035] | 1.010 (0.03) | [0.944,1.080] | 1.059 (0.06) | [0.950,1.180] |
| | Total peer-child relationship score, 2016 (Round 5) | 1.049 (0.04) | [0.971,1.133] | 1.041 (0.04) | [0.963,1.125] | 0.943 (0.04) | [0.868,1.026] | 1.001 (0.08) | [0.852,1.177] |
| | Middle/Top wealth tercile R5, 2016 (Round 5) | 0.732 (0.19) | [0.445,1.205] | 0.867 (0.22) | [0.522,1.440] | 2.247**(0.77) | [1.144,4.414] | 0.840 (0.27) | [0.445,1.585] |
| *Proxy baseline information* | | | | | | | | | |
| | Emotional problem scale (EPS) score, 2009 (Round 3) | 1.090* (0.05) | [0.995,1.195] | 1.101* (0.06) | [0.991,1.223] | 1.129**(0.07) | [1.003,1.270] | 1.114 (0.08) | [0.973,1.276] |
| | Subjective well-being, 2009 (Round 3) | 0.894 (0.06) | [0.782,1.023] | 1.077 (0.07) | [0.952,1.219] | 0.869* (0.07) | [0.736,1.026] | 0.952 (0.08) | [0.805,1.126] |
| | N | 650 | | 836 | | 391 | | 683 | |

*Note: Odds ratios are unadjusted odds ratios. Robust standard errors in parenthesis, *** significant at 1%, ** significant at 5%, * significant at 10%. Base categories are as follows: Male, Rural, Believe they are at no/low risk, Did not leave the house at all during the past 7 days, Did not spend more time taking care of children, Did not spend more time on household chores, Did not spend more time working in the family business, Did not face new health expenses, Did not suffer a shock, Did not work at all in the past 12 month OR worked during the pandemic but not before and is not working now, Does not have long-term health condition, Lowest wealth tercile. All time-variant variables are measured in 2020 unless otherwise specified. [3]In Vietnam only 17 people did not work before the pandemic but are working now. None showed depressive symptoms. As it predicts failure perfectly it drops out of the regressions.*

**Supplementary Table 10. Ethiopia: Means of continuous control variables by at least mild anxiety/depression**

| | | All | at least mild anxiety | no anxiety | p-value | at least mild depression | no depression | p-value |
|---|---|---|---|---|---|---|---|---|
| *Structural factors* | | | | | | | | |
| | Age in months | 250.86 | 264.14 | 247.97 | 0.000 | 255.55 | 250.00 | 0.014 |
| *COVID-19 related changes/behaviours* | | | | | | | | |
| | Difference in subjective well-being between Round 5 (2016) and phone survey (2020) | 0.96 | 1.15 | 0.92 | 0.052 | 1.26 | 0.91 | 0.005 |
| *Past protective/risk factors* | | | | | | | | |
| | Total parent-child relationship score, 2012/2016 (Round 4/5) | 26.26 | 25.98 | 26.32 | 0.036 | 26.20 | 26.27 | 0.686 |
| | Total peer-child relationship score, 2016 (Round 5) | 24.13 | 23.51 | 24.26 | 0.000 | 23.58 | 24.23 | 0.000 |
| *Proxy baseline information* | | | | | | | | |
| | Emotional problem scale (EPS) score, 2009 (Round 3)² | 2.78 | 3.00 | 2.70 | 0.173 | 3.13 | 2.71 | 0.096 |
| | Subjective well-being, 2009 (Round 3)² | 5.22 | 5.18 | 5.23 | 0.649 | 5.24 | 5.22 | 0.896 |

*Note*: *Results are for the combined Younger Cohort/ Older Cohort sample unless indicated otherwise, ² refers to Older Cohort (25-26) only.*

### Supplementary Table 11. India: Means of continuous control variables by at least mild anxiety/depression

| | | All | at least mild anxiety | no anxiety | p-value | at least mild depression | no depression | p-value |
|---|---|---|---|---|---|---|---|---|
| *Structural factors* | | | | | | | | |
| | Age in months | 253.30 | 261.16 | 252.32 | 0.000 | 258.12 | 252.77 | 0.037 |
| *COVID-19 related changes/behaviours* | | | | | | | | |
| | Difference in subjective well-being between Round 5 (2016) and phone survey (2020) | 0.49 | 0.77 | 0.45 | 0.009 | 0.86 | 0.45 | 0.001 |
| *Past protective/risk factors* | | | | | | | | |
| | Total parent-child relationship score, 2012/2016 (Round 4/5) | 27.63 | 26.94 | 27.72 | 0.000 | 26.84 | 27.72 | 0.000 |
| | Total peer-child relationship score, 2016 (Round 5) | 25.02 | 24.97 | 25.03 | 0.743 | 24.91 | 25.03 | 0.468 |
| *Proxy baseline information* | | | | | | | | |
| | Emotional problem scale (EPS) score, 2009 (Round 3)[2] | 3.57 | 3.93 | 3.51 | 0.067 | 4.14 | 3.49 | 0.010 |
| | Subjective well-being, 2009 (Round 3)[2] | 5.00 | 5.29 | 4.96 | 0.013 | 4.97 | 5.00 | 0.850 |

*Note*: *Results are for the combined Younger Cohort/ Older Cohort sample unless indicated otherwise, [2] refers to Older Cohort (25-26) only.*

**Supplementary Table 12. Peru: Means of continuous control variables by at least mild anxiety/depression**

| | | All | at least mild anxiety | no anxiety | p-value | at least mild depression | no depression | p-value |
|---|---|---|---|---|---|---|---|---|
| *Structural factors* | | | | | | | | |
| | Age in months | 244.23 | 245.19 | 243.57 | 0.315 | 242.56 | 245.00 | 0.151 |
| *COVID-19 related changes/behaviours* | | | | | | | | |
| | Difference in subjective well-being in Round 5 (2016) and phone survey (2020) | 0.52 | 0.74 | 0.38 | 0.000 | 0.70 | 0.44 | 0.013 |
| *Past protective/risk factors* | | | | | | | | |
| | Total parent-child relationship score, 2012/2016 (Round 4/5) | 24.86 | 24.60 | 25.04 | 0.009 | 24.28 | 25.12 | 0.000 |
| | Total peer-child relationship score, 2016 (Round 5) | 23.41 | 23.40 | 23.42 | 0.862 | 23.41 | 23.42 | 0.954 |
| *Proxy baseline information* | | | | | | | | |
| | Emotional problem scale (EPS) score, 2009 (Round 3)² | 4.27 | 4.87 | 3.81 | 0.000 | 4.85 | 4.03 | 0.002 |
| | Subjective well-being, 2009 (Round 3)² | 6.82 | 6.72 | 6.89 | 0.073 | 6.72 | 6.87 | 0.135 |

*Note*: *Results are for the combined Younger Cohort/ Older Cohort sample unless indicated otherwise, ² refers to Older Cohort (25-26) only.*

**Supplementary Table 13. Vietnam: Means of continuous control variables by at least mild anxiety/depression**

| | | All | at least mild anxiety | no anxiety | p-value | at least mild depression | no depression | p-value |
|---|---|---|---|---|---|---|---|---|
| *Structural factors* | | | | | | | | |
| | Age in months | 252.95 | 248.28 | 253.43 | 0.067 | 246.49 | 253.63 | 0.010 |
| *COVID-19 related changes/behaviours* | | | | | | | | |
| | Difference in subjective well-being in Round 5 (2016) and phone survey (2020) | -0.39 | 0.14 | -0.45 | 0.000 | 0.11 | -0.45 | 0.000 |
| *Past protective/risk factors* | | | | | | | | |
| | Total parent-child relationship score, 2012/2016 (Round 4/5) | 25.74 | 26.11 | 25.70 | 0.073 | 25.72 | 25.74 | 0.930 |
| | Total peer-child relationship score, 2016 (Round 5) | 22.57 | 22.40 | 22.59 | 0.230 | 22.52 | 22.58 | 0.704 |
| *Proxy baseline information* | | | | | | | | |
| | Emotional problem scale (EPS) score, 2009 (Round 3)[2] | 3.68 | 3.96 | 3.65 | 0.314 | 4.18 | 3.64 | 0.086 |
| | Subjective well-being, 2009 (Round 3)[2] | 5.84 | 5.73 | 5.86 | 0.383 | 5.85 | 5.84 | 0.943 |

*Note*: *Results are for the combined Younger Cohort/ Older Cohort sample unless indicated otherwise, [2] refers to Older Cohort (25-26) only.*

### Supplementary Table 14. Mental health outcomes by control variables (Ethiopia & India)

| | Ethiopia | | | | India | | | |
|---|---|---|---|---|---|---|---|---|
| | **% at least mild Anxiety** | p-value | **% at least mild Depression** | p-value | **% at least mild Anxiety** | p-value | **% at least mild Depression** | p-value |
| *COVID-19 related changes/behaviours* | | | | | | | | |
| Risk perception: believe they are at no/low risk | 20.30 | 0.048 | 20.75 | 0.000 | 10.10 | 0.092 | 10.10 | 0.735 |
| Risk perception: believe they are at medium/high risk | 16.79 | | 13.09 | | 12.17 | | 9.70 | |
| Did not leave house for any reason in the past 7 days | 22.50 | 0.173 | 18.33 | 0.367 | 8.46 | 0.030 | 5.51 | 0.000 |
| Left house for any reason in the past 7 days | 17.60 | | 15.27 | | 11.74 | | 11.07 | |
| *Change in responsibilities:* | | | | | | | | |
| Did not spend more time taking care of children | 18.06 | 0.704 | 15.55 | 0.823 | 9.01 | 0.000 | 8.82 | 0.000 |
| Spend more time taking care of children | 17.38 | | 15.17 | | 20.21 | | 14.79 | |
| Did not spend more time on household chores | 17.83 | 0.963 | 14.64 | 0.273 | 10.58 | 0.438 | 9.72 | 0.742 |
| Spend more time on household chores | 17.91 | | 16.34 | | 11.53 | | 10.10 | |
| Did not spend more time working in the family business | 18.26 | 0.209 | 15.63 | 0.516 | 10.90 | 0.204 | 9.90 | 0.924 |
| Spend more time working in the family business | 15.19 | | 14.13 | | 14.81 | | 10.19 | |
| *Economic shocks* | | | | | | | | |
| Was not faced with new health expenses | 17.43 | 0.115 | 15.75 | 0.230 | 10.91 | 0.827 | 8.57 | 0.041 |
| Faced with new health expenses | 21.72 | | 12.67 | | 11.18 | | 10.97 | |
| Did not experience adversity | 6.86 | 0.000 | 4.65 | 0.000 | 13.33 | 0.381 | 11.11 | 0.956 |
| Experienced adversity but did not reduce food consumption | 17.54 | | 15.59 | | 10.82 | | 9.88 | |
| Reduced food consumption as response to experienced adversity | 36.39 | | 31.29 | | 14.29 | | 10.20 | |
| *Changes in employment status* | | | | | | | | |
| Did not work at all in the past 12 month OR worked during the pandemic but not before the pandemic (and is not working now) | 13.79 | 0.000 | 13.10 | 0.000 | 9.22 | 0.000 | 9.03 | 0.059 |
| Did not work before the pandemic, but is working now | 26.17 | | 25.50 | | 9.07 | | 8.37 | |
| Worked before the pandemic and is working now/has a job | 18.34 | | 14.91 | | 12.89 | | 10.86 | |
| Worked before the pandemic and is not working now/does not have a job | 31.49 | | 22.65 | | 19.67 | | 15.57 | |
| *Educational disruption* | | | | | | | | |
| Never attended school or not enrolled in full-time education/not planning to enrol in learning activities[1] | 18.25 | 0.212 | 13.45 | 0.056 | 13.01 | 0.000 | 11.00 | 0.012 |
| Enrolled in/Planning to enrol in full time education and | 16.58 | | 17.41 | | 10.69 | | 10.40 | |

| | | | | | | | | |
|---|---|---|---|---|---|---|---|---|
| not participating in learning activities[1] | | | | | | | | |
| Enrolled in/Planning to enrol in full time education and participating in learning activities[1] | 21.13 | | 15.47 | | 6.57 | | 6.57 | |
| *Past protective/risk factors* | | | | | | | | |
| Participant does not have a long-term health problem, 2016 (Round 5) | 17.13 | 0.004 | 15.42 | 0.940 | 10.47 | 0.007 | 9.31 | 0.004 |
| Participant has long-term health problem, 2016 (Round 5) | 25.52 | | 15.63 | | 15.61 | | 14.62 | |

*Notes: If any missing answers to questions then the whole score is set to missing. p-value indicates significance of t-tests between dummy variables and o F-tests in case of categorical variables. Results are for the combined Younger Cohort/ Older Cohort sample unless indicated otherwise, [1] refers to Younger Cohort (18-19) only.*

### Supplementary Table 15. Mental health outcomes by control variables (Peru & Vietnam)

| | **Peru** | | | | **Vietnam** | | | |
|---|---|---|---|---|---|---|---|---|
| | **% at least mild Anxiety** | p-value | **% at least mild Depression** | p-value | **% at least mild Anxiety** | p-value | **% at least mild Depression** | p-value |
| *COVID-19 related changes/behaviours* | | | | | | | | |
| Risk perception: believe they are at no/low risk | 35.34 | 0.000 | 25.84 | 0.000 | 9.31 | 0.967 | 9.31 | 0.557 |
| Risk perception: believe they are at medium/high risk | 45.51 | | 36.43 | | 9.37 | | 10.18 | |
| Did not leave house for any reason in the past 7 days | 42.82 | 0.393 | 33.33 | 0.421 | 9.54 | 0.892 | 11.66 | 0.184 |
| Left house for any reason in the past 7 days | 40.38 | | 31.16 | | 9.29 | | 9.19 | |
| *Change in responsibilities:* | | | | | | | | |
| Did not spend more time taking care of children | 36.83 | 0.000 | 29.57 | 0.009 | 8.19 | 0.001 | 8.19 | 0.000 |
| Spend more time taking care of children | 48.61 | | 35.45 | | 13.12 | | 13.88 | |
| Did not spend more time on household chores | 33.96 | 0.001 | 28.77 | 0.157 | 7.44 | 0.003 | 6.46 | 0.000 |
| Spend more time on household chores | 42.86 | | 32.40 | | 11.09 | | 12.36 | |
| Did not spend more time working in the family business | 40.09 | 0.105 | 30.92 | 0.141 | 8.46 | 0.000 | 8.60 | 0.000 |
| Spend more time working in the family business | 45.23 | | 35.34 | | 16.40 | | 16.80 | |
| *Economic shocks* | | | | | | | | |
| Was not faced with new health expenses | 36.12 | 0.000 | 27.24 | 0.000 | 8.99 | 0.041 | 9.27 | 0.171 |
| Faced with new health expenses | 52.15 | | 41.94 | | 14.00 | | 12.67 | |

| | Peru | | | | Vietnam | | | |
|---|---|---|---|---|---|---|---|---|
| | **% at least mild Anxiety** | p-value | **% at least mild Depression** | p-value | **% at least mild Anxiety** | p-value | **% at least mild Depression** | p-value |
| Did not experience adversity | 16.33 | 0.002 | 14.29 | 0.004 | 6.49 | 0.000 | 6.29 | 0.000 |
| Experienced adversity but did not reduce food consumption | 41.32 | | 31.47 | | 10.80 | | 11.11 | |
| Reduced food consumption as response to experienced adversity | 44.26 | | 40.16 | | 12.95 | | 13.77 | |
| *Changes in employment status* | | | | | | | | |
| Did not work at all in the past 12 month OR worked during the pandemic but not before the pandemic (and is not working now) | 41.41 | 0.176 | 33.26 | 0.003 | 8.53 | 0.269 | 9.83 | 0.002 |
| Did not work before the pandemic, but is working now | 37.77 | | 32.19 | | 10.97 | | 8.39 | |
| Worked before the pandemic and is working now/has a job | 39.57 | | 27.89 | | 8.81 | | 8.11 | |
| Worked before the pandemic and is not working now/does not have a job | 45.91 | | 38.99 | | 12.01 | | 15.26 | |
| *Educational disruption* | | | | | | | | |
| Never attended school or not enrolled in full-time education/not planning to enrol in learning activities[1] | 40.74 | 0.611 | 30.74 | 0.490 | 9.95 | 0.374 | 8.91 | 0.387 |
| Enrolled in/Planning to enrol in full time education and not participating in learning activities[1] | 32.14 | | 28.57 | | 8.96 | | 9.43 | |
| Enrolled in/Planning to enrol in full time education and participating in learning activities[1] | 41.47 | | 33.33 | | 8.04 | | 10.82 | |
| *Past protective/risk factors* | | | | | | | | |
| Participant does not have a long-term health problem, 2016 (Round 5) | 38.71 | 0.000 | 29.85 | 0.000 | 9.04 | 0.248 | 9.14 | 0.151 |
| Participant has long-term health problem, 2016 (Round 5) | 55.65 | | 43.51 | | 11.04 | | 11.66 | |

*Notes: If any missing answers to questions then the whole score is set to missing. p-value indicates significance of t-tests between dummy variables and o F-tests in case of categorical variables. Results are for the combined Younger Cohort/ Older Cohort sample unless indicated otherwise, [1] refers to Younger Cohort (18-19) only.*

**Supplementary Table 16. Logistic regression results: Symptoms of at least mild anxiety/depression by gender (Ethiopia)**

| | At least mild anxiety | | | | At least mild depression | | | |
|---|---|---|---|---|---|---|---|---|
| | Male | | Female | | Male | | Female | |
| | Odds Ratio | 95% CI | Odds Ratio | 95% CI | Odds Ratio | 95% CI | Odds Ratio | 95% CI |
| *Structural factors* | | | | | | | | |
| Age in months | 1.012*** (0.00) | [1.007,1.016] | 1.003 (0.00) | [0.999,1.008] | 1.007*** (0.00) | [1.002,1.012] | 0.996* (0.00) | [0.991,1.001] |
| Urban | 1.155 (0.22) | [0.799,1.670] | 1.523** (0.32) | [1.003,2.312] | 1.307 (0.26) | [0.880,1.942] | 1.440* (0.31) | [0.938,2.210] |
| *COVID-19 related changes/behaviours* | | | | | | | | |
| Risk perception: believe they are at medium/high risk | 0.807 (0.14) | [0.569,1.146] | 0.825 (0.16) | [0.562,1.211] | 0.576*** (0.10) | [0.403,0.824] | 0.659** (0.13) | [0.452,0.961] |
| Left house for any reason in the past 7 days | 0.593 (0.29) | [0.229,1.533] | 0.847 (0.25) | [0.480,1.494] | 0.556 (0.25) | [0.233,1.327] | 1.126 (0.35) | [0.614,2.064] |
| Difference in subjective well-being between Round 5 (2016) and phone survey (2020) | 1.097** (0.04) | [1.016,1.185] | 1.052 (0.05) | [0.967,1.146] | 1.084* (0.05) | [0.999,1.177] | 1.114** (0.05) | [1.019,1.218] |
| *Change in responsibilities:* | | | | | | | | |
| Spend more time taking care of children | 1.262 (0.34) | [0.747,2.132] | 0.749 (0.14) | [0.513,1.093] | 0.791 (0.21) | [0.468,1.339] | 0.957 (0.20) | [0.640,1.433] |
| Spend more time on household chores | 0.869 (0.20) | [0.558,1.353] | 1.447 (0.33) | [0.929,2.252] | 1.181 (0.25) | [0.776,1.797] | 1.162 (0.27) | [0.731,1.845] |
| Spend more time working in the family business | 0.750 (0.20) | [0.448,1.256] | 0.713 (0.24) | [0.367,1.384] | 0.643 (0.19) | [0.361,1.144] | 1.498 (0.47) | [0.814,2.755] |
| *Economic shocks* | | | | | | | | |
| Faced with new health expenses | 0.733 (0.21) | [0.419,1.281] | 1.115 (0.30) | [0.660,1.886] | 0.438*** (0.13) | [0.241,0.797] | 0.592* (0.19) | [0.319,1.096] |
| Experienced adversity but did not reduce food consumption | 1.889** (0.53) | [1.091,3.270] | 3.063*** (1.00) | [1.619,5.793] | 3.658*** (1.35) | [1.776,7.537] | 3.739*** (1.28) | [1.908,7.326] |
| Reduced food consumption as response to experienced adversity | 5.891*** (1.92) | [3.107,11.172] | 8.642*** (3.16) | [4.222,17.690] | 12.987*** (5.25) | [5.884,28.663] | 8.585*** (3.32) | [4.026,18.308] |
| *Changes in employment status* | | | | | | | | |
| Did not work before the pandemic, but is working now | 2.623*** (0.88) | [1.356,5.075] | 2.853*** (0.97) | [1.470,5.538] | 2.426*** (0.79) | [1.283,4.591] | 2.754*** (0.99) | [1.357,5.590] |
| Worked before the pandemic and is working now/has a job | 1.335 (0.30) | [0.859,2.074] | 1.981*** (0.46) | [1.260,3.115] | 1.057 (0.23) | [0.684,1.634] | 1.744** (0.41) | [1.096,2.776] |

| | At least mild anxiety | | | | At least mild depression | | | |
|---|---|---|---|---|---|---|---|---|
| | Male | | Female | | Male | | Female | |
| | **Odds Ratio** | **95% CI** | **Odds Ratio** | **95% CI** | **Odds Ratio** | **95% CI** | **Odds Ratio** | **95% CI** |
| Worked before the pandemic and is not working now/does not have a job | 2.196*** (0.61) | [1.268,3.801] | 2.239*** (0.64) | [1.279,3.918] | 1.020 (0.33) | [0.540,1.927] | 2.647*** (0.79) | [1.476,4.746] |
| _Past protective/risk factors_ | | | | | | | | |
| Participant has long-term health problem, 2016 (Round 5) | 1.848** (0.53) | [1.054,3.241] | 1.204 (0.29) | [0.745,1.944] | 0.919 (0.33) | [0.450,1.875] | 0.998 (0.28) | [0.580,1.716] |
| Total parent-child relationship score, 2012/2016 (Round 4/5) | 0.968 (0.03) | [0.914,1.026] | 0.962 (0.03) | [0.902,1.026] | 1.027 (0.03) | [0.965,1.092] | 0.972 (0.03) | [0.908,1.039] |
| Total peer-child relationship score, 2016 (Round 5) | 0.935** (0.03) | [0.877,0.998] | 0.944* (0.03) | [0.890,1.002] | 0.948 (0.03) | [0.884,1.017] | 0.932** (0.03) | [0.875,0.993] |
| Middle/Top wealth tercile R5, 2016 (Round 5) | 1.199 (0.24) | [0.811,1.773] | 0.985 (0.21) | [0.643,1.511] | 0.882 (0.18) | [0.587,1.327] | 0.844 (0.19) | [0.539,1.321] |
| N | 1173 | | 1010 | | 1173 | | 1010 | |

*Note: Odds ratios are unadjusted odds ratios. Robust standard errors in parenthesis, *** significant at 1%, ** significant at 5%, * significant at 10%. Base categories are as follows: Rural, Believe they are at no/low risk, Did not leave the house at all during the past 7 days, Did not spend more time taking care of children, Did not spend more time on household chores, Did not spend more time working in the family business, Did not face new health expenses, Did not suffer a shock, Did not work at all in the past 12 month OR worked during the pandemic but not before and is not working now, Does not have long-term health condition, Lowest wealth tercile. All time-variant variables are measured in 2020 unless otherwise specified. Results are for the combined Younger Cohort/ Older Cohort sample.*

### Supplementary Table 17. Logistic regression results: Symptoms of at least mild anxiety/depression by gender (India)

| | At least mild anxiety | | | | At least mild depression | | | |
|---|---|---|---|---|---|---|---|---|
| | Male | | Female | | Male | | Female | |
| | **Odds Ratio** | **95% CI** | **Odds Ratio** | **95% CI** | **Odds Ratio** | **95% CI** | **Odds Ratio** | **95% CI** |
| _Structural factors_ | | | | | | | | |
| Age in months | 0.999 (0.00) | [0.994,1.004] | 1.003 (0.00) | [0.998,1.008] | 0.998 (0.00) | [0.993,1.003] | 1.002 (0.00) | [0.996,1.008] |
| Urban | 0.428*** (0.12) | [0.252,0.728] | 1.564** (0.35) | [1.012,2.417] | 0.502*** (0.12) | [0.311,0.808] | 1.113 (0.28) | [0.679,1.824] |
| _COVID-19 related changes/behaviours_ | | | | | | | | |
| Risk perception: believe they are at medium/high risk | 1.520** (0.30) | [1.038,2.224] | 1.149 (0.22) | [0.795,1.661] | 1.048 (0.20) | [0.722,1.522] | 0.927 (0.19) | [0.616,1.395] |
| Left house for any reason in the past 7 days | 0.744 (0.28) | [0.356,1.555] | 1.606** (0.36) | [1.040,2.482] | 1.063 (0.43) | [0.482,2.342] | 2.810*** (0.77) | [1.642,4.810] |

| | At least mild anxiety | | | | At least mild depression | | | |
|---|---|---|---|---|---|---|---|---|
| | Male | | Female | | Male | | Female | |
| | Odds Ratio | 95% CI | Odds Ratio | 95% CI | Odds Ratio | 95% CI | Odds Ratio | 95% CI |
| Difference in subjective well-being between Round 5 (2016) and phone survey (2020) | 0.999 (0.05) | [0.904,1.105] | 1.245*** (0.07) | [1.121,1.383] | 1.041 (0.05) | [0.950,1.141] | 1.248*** (0.07) | [1.117,1.395] |
| *Change in responsibilities:* | | | | | | | | |
| Spend more time taking care of children | 1.708* (0.50) | [0.967,3.015] | 2.455*** (0.54) | [1.595,3.780] | 1.327 (0.41) | [0.721,2.443] | 1.668** (0.43) | [1.007,2.761] |
| Spend more time on household chores | 1.356 (0.27) | [0.923,1.993] | 0.440*** (0.09) | [0.297,0.651] | 1.337 (0.26) | [0.915,1.955] | 0.576** (0.13) | [0.376,0.883] |
| Spend more time working in the family business | 1.944* (0.71) | [0.953,3.967] | 1.210 (0.52) | [0.520,2.816] | 1.100 (0.45) | [0.493,2.457] | 1.043 (0.58) | [0.348,3.122] |
| *Economic shocks* | | | | | | | | |
| Faced with new health expenses | 0.786 (0.16) | [0.532,1.161] | 1.349 (0.26) | [0.924,1.968] | 1.203 (0.25) | [0.799,1.810] | 1.477* (0.32) | [0.969,2.250] |
| Experienced adversity but did not reduce food consumption | 0.894 (0.76) | [0.169,4.723] | 0.500 (0.30) | [0.155,1.609] | 1.749 (1.91) | [0.207,14.799] | 0.384* (0.21) | [0.130,1.137] |
| Reduced food consumption as response to experienced adversity | 1.272 (1.17) | [0.209,7.752] | 0.691 (0.48) | [0.177,2.697] | 1.651 (1.92) | [0.169,16.089] | 0.398 (0.28) | [0.099,1.608] |
| *Changes in employment status* | | | | | | | | |
| Did not work before the pandemic, but is working now | 0.629 (0.21) | [0.326,1.213] | 1.267 (0.40) | [0.687,2.334] | 0.555* (0.18) | [0.294,1.051] | 0.887 (0.29) | [0.472,1.669] |
| Worked before the pandemic and is working now/has a job | 0.982 (0.28) | [0.562,1.718] | 1.504* (0.33) | [0.978,2.314] | 0.980 (0.25) | [0.591,1.626] | 0.802 (0.21) | [0.485,1.328] |
| Worked before the pandemic and is not working now/does not have a job | 2.812** (1.14) | [1.266,6.245] | 2.286** (0.83) | [1.121,4.660] | 2.908*** (1.12) | [1.367,6.188] | 0.836 (0.40) | [0.325,2.148] |
| *Past protective/risk factors* | | | | | | | | |
| Participant has long-term health problem, 2016 (Round 5) | 1.310 (0.38) | [0.746,2.299] | 1.466 (0.35) | [0.924,2.326] | 1.353 (0.39) | [0.773,2.366] | 1.785** (0.44) | [1.105,2.885] |
| Total parent-child relationship score, 2012/2016 (Round 4/5) | 0.908*** (0.03) | [0.845,0.977] | 0.934** (0.03) | [0.880,0.991] | 0.908*** (0.03) | [0.846,0.973] | 0.909*** (0.03) | [0.848,0.974] |
| Total peer-child relationship score, 2016 (Round 5) | 1.039 (0.04) | [0.965,1.119] | 1.014 (0.04) | [0.944,1.089] | 1.040 (0.04) | [0.961,1.125] | 0.994 (0.04) | [0.924,1.069] |
| Middle/Top wealth tercile R5, 2016 (Round 5) | 0.804 (0.17) | [0.534,1.210] | 0.764 (0.16) | [0.504,1.159] | 1.324 (0.27) | [0.881,1.989] | 0.877 (0.20) | [0.559,1.375] |
| N | 1369 | | 1253 | | 1369 | | 1253 | |

*Note: Odds ratios are unadjusted odds ratios. Robust standard errors in parenthesis, *** significant at 1%, ** significant at 5%, * significant at 10%. Base categories are as follows: Rural, Believe they are at no/low risk, Did not leave the house at all during the past 7 days, Did not spend more time taking care of children, Did not spend more time on household chores, Did not spend more time working in the family business, Did not face new health expenses, Did not suffer a shock, Did not work at all in the past 12 month OR worked during the pandemic but not before and is not working now, Does not have long-term health condition, Lowest wealth tercile. All time-variant variables are measured in 2020 unless otherwise specified. Results are for the combined Younger Cohort/ Older Cohort sample.*

**Supplementary Table 18. Logistic regression results: Symptoms of at least mild anxiety/depression by gender (Peru)**

| | At least mild anxiety | | | | At least mild depression | | | |
|---|---|---|---|---|---|---|---|---|
| | Male | | Female | | Male | | Female | |
| | Odds Ratio | 95% CI | Odds Ratio | 95% CI | Odds Ratio | 95% CI | Odds Ratio | 95% CI |
| *Structural factors* | | | | | | | | |
| Age in months | 1.001 (0.00) | [0.997,1.005] | 0.998 (0.00) | [0.994,1.002] | 0.996* (0.00) | [0.991,1.001] | 0.996* (0.00) | [0.992,1.000] |
| Urban | 1.245 (0.27) | [0.817,1.898] | 1.610** (0.34) | [1.070,2.423] | 0.978 (0.21) | [0.639,1.498] | 1.222 (0.27) | [0.790,1.889] |
| *COVID-19 related changes/behaviours* | | | | | | | | |
| Risk perception: believe they are at medium/high risk | 1.326* (0.19) | [0.994,1.767] | 1.585*** (0.22) | [1.204,2.086] | 1.513*** (0.23) | [1.117,2.051] | 1.657*** (0.24) | [1.241,2.212] |
| Left house for any reason in the past 7 days | 1.324 (0.28) | [0.878,1.996] | 0.848 (0.14) | [0.609,1.181] | 1.052 (0.22) | [0.697,1.588] | 1.067 (0.19) | [0.756,1.505] |
| Difference in subjective well-being between Round 5 (2016) and phone survey (2020) | 1.182*** (0.05) | [1.097,1.274] | 1.064* (0.03) | [0.998,1.134] | 1.131*** (0.05) | [1.043,1.227] | 1.047 (0.04) | [0.978,1.121] |
| *Change in responsibilities:* | | | | | | | | |
| Spend more time taking care of children | 1.567*** (0.26) | [1.134,2.165] | 1.285* (0.18) | [0.973,1.698] | 1.529** (0.26) | [1.097,2.131] | 0.932 (0.14) | [0.697,1.246] |
| Spend more time on household chores | 1.220 (0.21) | [0.873,1.704] | 1.293 (0.25) | [0.882,1.895] | 1.039 (0.18) | [0.734,1.471] | 1.105 (0.22) | [0.742,1.644] |
| Spend more time working in the family business | 1.299 (0.25) | [0.896,1.885] | 1.335 (0.29) | [0.871,2.046] | 1.258 (0.25) | [0.848,1.865] | 1.362 (0.30) | [0.883,2.101] |
| *Economic shocks* | | | | | | | | |
| Faced with new health expenses | 1.983*** (0.32) | [1.451,2.710] | 1.532*** (0.23) | [1.148,2.044] | 1.735*** (0.29) | [1.257,2.394] | 1.757*** (0.26) | [1.308,2.361] |
| Experienced adversity but did not reduce food consumption | 5.087** (3.94) | [1.114,23.224] | 1.609 (0.88) | [0.548,4.728] | 4.172* (3.08) | [0.983,17.705] | 1.056 (0.63) | [0.330,3.378] |
| Reduced food consumption as response to experienced adversity | 5.843** (4.83) | [1.157,29.524] | 1.288 (0.77) | [0.397,4.178] | 5.041** (4.02) | [1.057,24.045] | 1.354 (0.87) | [0.383,4.782] |
| *Changes in employment status* | | | | | | | | |
| Did not work before the pandemic, but is working now | 0.684 (0.21) | [0.379,1.235] | 0.945 (0.23) | [0.591,1.510] | 1.065 (0.32) | [0.593,1.910] | 0.790 (0.19) | [0.491,1.271] |
| Worked before the pandemic and is working now/has a job | 0.958 (0.22) | [0.608,1.509] | 1.049 (0.19) | [0.737,1.493] | 0.899 (0.22) | [0.563,1.437] | 0.799 (0.15) | [0.551,1.158] |
| Worked before the pandemic and is not working now/does not have a job | 1.029 (0.27) | [0.616,1.719] | 1.326 (0.26) | [0.900,1.952] | 1.385 (0.37) | [0.824,2.329] | 1.193 (0.25) | [0.797,1.786] |

| | At least mild anxiety | | | | At least mild depression | | | |
|---|---|---|---|---|---|---|---|---|
| | Male | | Female | | Male | | Female | |
| | Odds Ratio | 95% CI | Odds Ratio | 95% CI | Odds Ratio | 95% CI | Odds Ratio | 95% CI |
| *Past protective/risk factors* | | | | | | | | |
| Participant has long-term health problem, 2016 (Round 5) | 1.794** (0.41) | [1.142,2.818] | 1.937*** (0.38) | [1.318,2.848] | 1.662** (0.39) | [1.047,2.640] | 1.564** (0.29) | [1.081,2.262] |
| Total parent-child relationship score, 2012/2016 (Round 4/5) | 0.974 (0.02) | [0.931,1.020] | 0.982 (0.02) | [0.947,1.019] | 0.944** (0.02) | [0.898,0.992] | 0.936*** (0.02) | [0.900,0.973] |
| Total peer-child relationship score, 2016 (Round 5) | 1.010 (0.03) | [0.958,1.065] | 0.990 (0.02) | [0.951,1.031] | 1.002 (0.03) | [0.946,1.063] | 1.011 (0.02) | [0.968,1.055] |
| Middle/Top wealth tercile R5, 2016 (Round 5) | 0.777 (0.14) | [0.548,1.104] | 1.257 (0.22) | [0.890,1.774] | 1.137 (0.21) | [0.795,1.627] | 1.294 (0.24) | [0.894,1.873] |
| N | 957 | | 930 | | 957 | | 930 | |

*Note: Odds ratios are unadjusted odds ratios. Robust standard errors in parenthesis, *** significant at 1%, ** significant at 5%, * significant at 10%. Base categories are as follows: Rural, Believe they are at no/low risk, Did not leave the house at all during the past 7 days, Did not spend more time taking care of children, Did not spend more time on household chores, Did not spend more time working in the family business, Did not face new health expenses, Did not suffer a shock, Did not work at all in the past 12 month OR worked during the pandemic but not before and is not working now, Does not have long-term health condition, Lowest wealth tercile. All time-variant variables are measured in 2020 unless otherwise specified. Results are for the combined Younger Cohort/ Older Cohort sample.*

### Supplementary Table 19. Logistic regression results: Symptoms of at least mild anxiety/depression by gender (Vietnam)

| | At least mild anxiety | | | | At least mild depression | | | |
|---|---|---|---|---|---|---|---|---|
| | Male | | Female | | Male | | Female | |
| | Odds Ratio | 95% CI | Odds Ratio | 95% CI | Odds Ratio | 95% CI | Odds Ratio | 95% CI |
| *Structural factors* | | | | | | | | |
| Age in months | 0.996 (0.00) | [0.990,1.002] | 0.994** (0.00) | [0.988,1.000] | 0.992** (0.00) | [0.985,0.999] | 0.994** (0.00) | [0.988,1.000] |
| Urban | 0.960 (0.22) | [0.608,1.517] | 1.358 (0.27) | [0.916,2.014] | 0.837 (0.22) | [0.500,1.402] | 1.091 (0.22) | [0.740,1.607] |
| *COVID-19 related changes/behaviours* | | | | | | | | |
| Risk perception: believe they are at medium/high risk | 1.548* (0.41) | [0.921,2.603] | 0.816 (0.21) | [0.498,1.337] | 2.094** (0.61) | [1.180,3.715] | 0.828 (0.19) | [0.525,1.304] |
| Left house for any reason in the past 7 days | 0.666 (0.22) | [0.353,1.254] | 1.535 (0.51) | [0.802,2.936] | 0.748 (0.25) | [0.388,1.439] | 1.225 (0.36) | [0.694,2.164] |
| Difference in subjective well-being between | 1.296*** (0.10) | [1.116,1.506] | 1.111* (0.06) | [0.994,1.241] | 1.471*** (0.11) | [1.269,1.706] | 1.016 (0.05) | [0.914,1.129] |

| | At least mild anxiety | | | | At least mild depression | | | |
|---|---|---|---|---|---|---|---|---|
| | Male | | Female | | Male | | Female | |
| | **Odds Ratio** | **95% CI** | **Odds Ratio** | **95% CI** | **Odds Ratio** | **95% CI** | **Odds Ratio** | **95% CI** |
| Round 5 (2016) and phone survey (2020) | | | | | | | | |
| *Change in responsibilities:* | | | | | | | | |
| Spend more time taking care of children | 1.222 (0.37) | [0.673,2.220] | 1.520* (0.35) | [0.971,2.381] | 1.221 (0.39) | [0.648,2.300] | 1.506* (0.34) | [0.970,2.339] |
| Spend more time on household chores | 1.191 (0.27) | [0.757,1.872] | 1.191 (0.26) | [0.777,1.825] | 1.076 (0.27) | [0.655,1.770] | 1.996*** (0.46) | [1.277,3.122] |
| Spend more time working in the family business | 2.394*** (0.75) | [1.292,4.434] | 1.624* (0.44) | [0.950,2.777] | 2.373*** (0.76) | [1.261,4.463] | 1.875** (0.48) | [1.132,3.106] |
| *Economic shocks* | | | | | | | | |
| Faced with new health expenses | 0.544 (0.29) | [0.191,1.547] | 1.757* (0.57) | [0.933,3.309] | 1.093 (0.49) | [0.453,2.636] | 0.721 (0.27) | [0.345,1.508] |
| Experienced adversity but did not reduce food consumption | 2.494*** (0.70) | [1.440,4.320] | 1.230 (0.28) | [0.787,1.925] | 2.430*** (0.71) | [1.370,4.309] | 1.416 (0.33) | [0.902,2.224] |
| Reduced food consumption as response to experienced adversity | 2.367** (0.86) | [1.159,4.833] | 1.357 (0.38) | [0.785,2.346] | 1.883* (0.72) | [0.889,3.987] | 1.946** (0.51) | [1.162,3.259] |
| *Changes in employment status* | | | | | | | | |
| Did not work before the pandemic, but is working now | 1.605 (0.84) | [0.574,4.486] | 0.814 (0.34) | [0.357,1.857] | 0.254 (0.25) | [0.036,1.782] | 0.913 (0.34) | [0.436,1.910] |
| Worked before the pandemic and is working now/has a job | 1.127 (0.42) | [0.545,2.331] | 0.948 (0.24) | [0.575,1.565] | 1.157 (0.41) | [0.576,2.324] | 0.708 (0.17) | [0.441,1.136] |
| Worked before the pandemic and is not working now/does not have a job | 1.798 (0.72) | [0.817,3.958] | 1.178 (0.37) | [0.635,2.187] | 1.921* (0.75) | [0.898,4.108] | 1.456 (0.40) | [0.848,2.502] |
| *Past protective/risk factors* | | | | | | | | |
| Participant has long-term health problem, 2016 (Round 5) | 1.944** (0.58) | [1.088,3.474] | 1.055 (0.29) | [0.612,1.819] | 2.231** (0.74) | [1.167,4.266] | 1.063 (0.28) | [0.631,1.789] |
| Total parent-child relationship score, 2012/2016 (Round 4/5) | 1.015 (0.04) | [0.940,1.096] | 1.056 (0.04) | [0.989,1.127] | 0.983 (0.04) | [0.904,1.071] | 0.982 (0.03) | [0.925,1.043] |
| Total peer-child relationship score, 2016 | 1.014 (0.06) | [0.907,1.133] | 0.897** (0.04) | [0.814,0.989] | 1.055 (0.07) | [0.928,1.199] | 0.961 (0.04) | [0.877,1.052] |

| | At least mild anxiety | | | | At least mild depression | | | |
|---|---|---|---|---|---|---|---|---|
| | Male | | Female | | Male | | Female | |
| | **Odds Ratio** | **95% CI** | **Odds Ratio** | **95% CI** | **Odds Ratio** | **95% CI** | **Odds Ratio** | **95% CI** |
| (Round 5) | | | | | | | | |
| Middle/Top wealth tercile R5, 2016 (Round 5) | 0.551** (0.14) | [0.336,0.903] | 0.566*** (0.12) | [0.379,0.844] | 1.015 (0.28) | [0.588,1.754] | 1.130 (0.24) | [0.745,1.713] |
| N | 1111 | | 1185 | | 1111 | | 1185 | |

*Note: Odds ratios are unadjusted odds ratios. Robust standard errors in parenthesis, *** significant at 1%, ** significant at 5%, * significant at 10%. Base categories are as follows: Rural, Believe they are at no/low risk, Did not leave the house at all during the past 7 days, Did not spend more time taking care of children, Did not spend more time on household chores, Did not spend more time working in the family business, Did not face new health expenses, Did not suffer a shock, Did not work at all in the past 12 month OR worked during the pandemic but not before and is not working now, Does not have long-term health condition, Lowest wealth tercile. All time-variant variables are measured in 2020 unless otherwise specified. Results are for the combined Younger Cohort/ Older Cohort sample.*

**Supplementary Table 20. Logistic regression results: Symptoms of at least mild anxiety/depression by gender for the Younger Cohort only (Ethiopia)**

| | At least mild anxiety | | | | At least mild depression | | | |
|---|---|---|---|---|---|---|---|---|
| | Male | | Female | | Male | | Female | |
| | Odds Ratio | 95% CI | Odds Ratio | 95% CI | Odds Ratio | 95% CI | Odds Ratio | 95% CI |
| *Structural factors* | | | | | | | | |
| Age in months | 1.011 (0.03) | [0.952,1.074] | 0.990 (0.03) | [0.937,1.047] | 1.005 (0.03) | [0.946,1.068] | 0.997 (0.03) | [0.944,1.054] |
| Urban | 1.421 (0.39) | [0.831,2.430] | 1.712* (0.51) | [0.953,3.075] | 1.341 (0.37) | [0.783,2.296] | 1.634* (0.45) | [0.956,2.792] |
| *COVID-19 related changes/behaviours* | | | | | | | | |
| Risk perception: believe they are at medium/high risk | 0.550** (0.13) | [0.348,0.869] | 0.875 (0.22) | [0.540,1.420] | 0.461*** (0.11) | [0.290,0.733] | 0.512*** (0.12) | [0.324,0.810] |
| Left house for any reason in the past 7 days | 0.620 (0.29) | [0.249,1.545] | 1.140 (0.47) | [0.504,2.579] | 0.702 (0.34) | [0.269,1.832] | 1.129 (0.42) | [0.546,2.336] |
| Difference in subjective well-being between Round 5 (2016) and phone survey (2020) | 1.076 (0.05) | [0.973,1.189] | 1.001 (0.06) | [0.897,1.117] | 1.015 (0.06) | [0.910,1.131] | 1.083 (0.06) | [0.975,1.204] |
| *Change in responsibilities:* | | | | | | | | |
| Spend more time taking care of children | 1.481 (0.44) | [0.823,2.666] | 0.754 (0.19) | [0.460,1.236] | 0.839 (0.25) | [0.463,1.522] | 1.224 (0.30) | [0.757,1.979] |
| Spend more time on household chores | 0.780 (0.19) | [0.479,1.271] | 1.401 (0.41) | [0.793,2.475] | 1.083 (0.26) | [0.671,1.749] | 1.132 (0.32) | [0.647,1.979] |
| Spend more time working in the family business | 0.542 (0.23) | [0.239,1.233] | 0.842 (0.39) | [0.344,2.064] | 0.475 (0.22) | [0.189,1.195] | 2.336** (0.88) | [1.120,4.872] |
| *Economic shocks* | | | | | | | | |
| Faced with new health expenses | 0.673 (0.31) | [0.272,1.666] | 0.645 (0.28) | [0.273,1.524] | 0.278** (0.17) | [0.083,0.931] | 0.199** (0.13) | [0.057,0.695] |
| Experienced adversity but did not reduce food consumption | 3.346*** (1.34) | [1.530,7.320] | 3.633*** (1.42) | [1.692,7.801] | 6.595*** (3.49) | [2.338,18.604] | 3.859*** (1.52) | [1.785,8.340] |
| Reduced food consumption as response to experienced adversity | 6.188*** (2.97) | [2.418,15.835] | 9.701*** (4.34) | [4.035,23.323] | 19.255*** (11.13) | [6.202,59.780] | 8.538*** (3.98) | [3.427,21.271] |
| *Educational disruption* | | | | | | | | |
| Enrolled in/Planning to enrol in full time education and not participating in learning activities | 1.188 (0.33) | [0.693,2.039] | 2.556*** (0.87) | [1.313,4.972] | 1.520 (0.45) | [0.845,2.732] | 2.033** (0.62) | [1.122,3.683] |
| Enrolled in/Planning to enrol in full time education and participating in learning activities | 0.810 (0.46) | [0.269,2.436] | 1.600 (0.79) | [0.608,4.207] | 0.735 (0.48) | [0.207,2.613] | 1.524 (0.68) | [0.635,3.656] |
| *Past protective/risk factors* | | | | | | | | |
| Participant has long-term health problem, | 1.185 | [0.464,3.028] | 1.291 | [0.630,2.646] | 0.358 | [0.086,1.494] | 0.621 | [0.260,1.483] |

| | At least mild anxiety | | | | At least mild depression | | | |
|---|---|---|---|---|---|---|---|---|
| | Male | | Female | | Male | | Female | |
| | Odds Ratio | 95% CI | Odds Ratio | 95% CI | Odds Ratio | 95% CI | Odds Ratio | 95% CI |
| 2016 (Round 5) | (0.57) | | (0.47) | | (0.26) | | (0.28) | |
| Total parent-child relationship score, 2012/2016 (Round 4/5) | 0.971 (0.04) | [0.890,1.059] | 0.943 (0.04) | [0.858,1.035] | 1.056 (0.04) | [0.974,1.145] | 0.970 (0.04) | [0.889,1.059] |
| Total peer-child relationship score, 2016 (Round 5) | 0.934 (0.05) | [0.848,1.028] | 0.970 (0.04) | [0.893,1.054] | 0.892** (0.05) | [0.808,0.985] | 0.915** (0.04) | [0.842,0.994] |
| Middle/Top wealth tercile R5, 2016 (Round 5) | 0.922 (0.26) | [0.528,1.611] | 0.923 (0.28) | [0.507,1.682] | 0.792 (0.23) | [0.453,1.383] | 0.756 (0.21) | [0.434,1.315] |
| N | 825 | | 708 | | 825 | | 708 | |

*Note: Odds ratios are unadjusted odds ratios. Robust standard errors in parenthesis, *** significant at 1%, ** significant at 5%, * significant at 10%. Base categories are as follows: Rural, Believe they are at no/low risk, Did not leave the house at all during the past 7 days, Did not spend more time taking care of children, Did not spend more time on household chores, Did not spend more time working in the family business, Did not face new health expenses, Did not suffer a shock, Never attended school or not enrolled in full-time education/not planning to enrol, Does not have long-term health condition, Lowest wealth tercile. All time-variant variables are measured in 2020 unless otherwise specified.*

### Supplementary Table 21. Logistic regression results: Symptoms of at least mild anxiety/depression by gender for the Younger Cohort only (India)

| | At least mild anxiety | | | | At least mild depression | | | |
|---|---|---|---|---|---|---|---|---|
| | Male | | Female | | Male | | Female | |
| | Odds Ratio | 95% CI | Odds Ratio | 95% CI | Odds Ratio | 95% CI | Odds Ratio | 95% CI |
| *Structural factors* | | | | | | | | |
| Age in months | 1.007 (0.03) | [0.947,1.069] | 0.971 (0.03) | [0.911,1.034] | 1.013 (0.03) | [0.955,1.075] | 0.968 (0.03) | [0.908,1.031] |
| Urban | 0.570* (0.18) | [0.303,1.070] | 2.637*** (0.74) | [1.517,4.585] | 0.769 (0.22) | [0.439,1.349] | 1.367 (0.45) | [0.720,2.594] |
| *COVID-19 related changes/behaviours* | | | | | | | | |
| Risk perception: believe they are at medium/high risk | 1.455 (0.34) | [0.914,2.315] | 0.995 (0.25) | [0.605,1.639] | 0.969 (0.22) | [0.622,1.509] | 0.989 (0.26) | [0.586,1.669] |
| Left house for any reason in the past 7 days | 0.517* (0.21) | [0.238,1.125] | 2.471*** (0.77) | [1.337,4.565] | 0.913 (0.39) | [0.392,2.124] | 3.923*** (1.55) | [1.811,8.496] |
| Difference in subjective well-being between Round 5 (2016) and phone survey (2020) | 1.012 (0.06) | [0.898,1.141] | 1.243*** (0.09) | [1.083,1.427] | 1.027 (0.06) | [0.919,1.148] | 1.215*** (0.09) | [1.058,1.395] |
| *Change in responsibilities:* | | | | | | | | |
| Spend more time taking care of children | 2.137* | [0.935,4.884] | 2.311** | [1.213,4.403] | 1.118 | [0.407,3.074] | 3.427*** | [1.798,6.534] |

| | At least mild anxiety | | | | At least mild depression | | | |
|---|---|---|---|---|---|---|---|---|
| | Male | | Female | | Male | | Female | |
| | Odds Ratio | 95% CI | Odds Ratio | 95% CI | Odds Ratio | 95% CI | Odds Ratio | 95% CI |
| | (0.90) | | (0.76) | | (0.58) | | (1.13) | |
| Spend more time on household chores | 1.370 (0.35) | [0.826,2.274] | 0.517*** (0.13) | [0.317,0.843] | 1.207 (0.30) | [0.738,1.973] | 0.673 (0.19) | [0.390,1.162] |
| Spend more time working in the family business | 2.020* (0.86) | [0.879,4.640] | 2.424** (1.09) | [1.005,5.849] | 0.750 (0.38) | [0.278,2.021] | 0.832 (0.58) | [0.212,3.262] |
| *Economic shocks* | | | | | | | | |
| Faced with new health expenses | 0.670 (0.17) | [0.413,1.088] | 1.165 (0.30) | [0.699,1.944] | 1.037 (0.25) | [0.645,1.667] | 1.171 (0.36) | [0.646,2.121] |
| Experienced adversity but did not reduce food consumption | 0.627 (0.52) | [0.125,3.157] | 0.691 (0.53) | [0.152,3.137] | 1.449 (1.56) | [0.177,11.888] | 0.313* (0.21) | [0.085,1.153] |
| Reduced food consumption as response to experienced adversity | 1.181 (1.09) | [0.193,7.208] | 0.882 (0.78) | [0.155,5.013] | 1.943 (2.25) | [0.201,18.787] | 0.481 (0.42) | [0.087,2.668] |
| *Educational disruption* | | | | | | | | |
| Enrolled in/Planning to enrol in full time education and not participating in learning activities | 0.934 (0.27) | [0.530,1.644] | 1.134 (0.34) | [0.628,2.049] | 0.914 (0.26) | [0.520,1.607] | 1.456 (0.47) | [0.770,2.753] |
| Enrolled in/Planning to enrol in full time education and participating in learning activities | 0.684 (0.24) | [0.342,1.369] | 0.659 (0.23) | [0.328,1.324] | 0.540* (0.19) | [0.276,1.057] | 1.122 (0.42) | [0.540,2.332] |
| <u>*Past protective/risk factors*</u> | | | | | | | | |
| Participant has long-term health problem, 2016 (Round 5) | 1.336 (0.46) | [0.676,2.640] | 1.199 (0.46) | [0.568,2.530] | 1.447 (0.49) | [0.748,2.802] | 1.429 (0.53) | [0.692,2.951] |
| Total parent-child relationship score, 2012/2016 (Round 4/5) | 0.918* (0.04) | [0.838,1.007] | 0.922* (0.04) | [0.846,1.005] | 0.921* (0.04) | [0.847,1.001] | 0.864*** (0.04) | [0.787,0.948] |
| Total peer-child relationship score, 2016 (Round 5) | 0.998 (0.05) | [0.908,1.096] | 1.011 (0.05) | [0.910,1.123] | 0.973 (0.05) | [0.877,1.080] | 1.040 (0.06) | [0.934,1.157] |
| Middle/Top wealth tercile R5, 2016 (Round 5) | 0.877 (0.23) | [0.524,1.468] | 0.586* (0.17) | [0.333,1.033] | 1.494 (0.41) | [0.877,2.546] | 1.129 (0.34) | [0.627,2.033] |
| N | 958 | | 828 | | 958 | | 828 | |

*Note: Odds ratios are unadjusted odds ratios. Robust standard errors in parenthesis, *** significant at 1%, ** significant at 5%, * significant at 10%. Base categories are as follows: Rural, Believe they are at no/low risk, Did not leave the house at all during the past 7 days, Did not spend more time taking care of children, Did not spend more time on household chores, Did not spend more time working in the family business, Did not face new health expenses, Did not suffer a shock, Never attended school or not enrolled in full-time education/not planning to enrol, Does not have long-term health condition, Lowest wealth tercile. All time-variant variables are measured in 2020 unless otherwise specified.*

**Supplementary Table 22. Logistic regression results: Symptoms of at least mild anxiety/depression by gender for the Younger Cohort only (Peru)**

| | At least mild anxiety | | | | At least mild depression | | | |
|---|---|---|---|---|---|---|---|---|
| | Male | | Female | | Male | | Female | |
| | **Odds Ratio** | **95% CI** | **Odds Ratio** | **95% CI** | **Odds Ratio** | **95% CI** | **Odds Ratio** | **95% CI** |
| *Structural factors* | | | | | | | | |
| Age in months | 0.981 (0.02) | [0.939,1.024] | 1.004 (0.02) | [0.965,1.045] | 0.953** (0.02) | [0.910,0.997] | 0.991 (0.02) | [0.952,1.033] |
| Urban | 1.090 (0.26) | [0.684,1.736] | 1.658** (0.37) | [1.070,2.569] | 1.043 (0.25) | [0.651,1.670] | 1.268 (0.30) | [0.793,2.029] |
| *COVID-19 related changes/behaviours* | | | | | | | | |
| Risk perception: believe they are at medium/high risk | 1.375* (0.23) | [0.988,1.913] | 1.488*** (0.23) | [1.100,2.014] | 1.500** (0.26) | [1.064,2.114] | 1.621*** (0.26) | [1.180,2.226] |
| Left house for any reason in the past 7 days | 1.228 (0.27) | [0.804,1.876] | 0.931 (0.17) | [0.650,1.332] | 0.998 (0.22) | [0.645,1.546] | 0.888 (0.17) | [0.614,1.284] |
| Difference in subjective well-being between Round 5 (2016) and phone survey (2020) | 1.190*** (0.05) | [1.097,1.291] | 1.060* (0.04) | [0.989,1.135] | 1.137*** (0.05) | [1.036,1.247] | 1.045 (0.04) | [0.970,1.126] |
| *Change in responsibilities:* | | | | | | | | |
| Spend more time taking care of children | 1.784*** (0.33) | [1.238,2.571] | 1.384** (0.22) | [1.013,1.890] | 1.823*** (0.35) | [1.247,2.665] | 0.965 (0.16) | [0.699,1.331] |
| Spend more time on household chores | 1.233 (0.25) | [0.832,1.826] | 1.176 (0.25) | [0.771,1.792] | 1.118 (0.23) | [0.748,1.671] | 1.121 (0.24) | [0.731,1.720] |
| Spend more time working in the family business | 1.094 (0.24) | [0.712,1.681] | 1.426 (0.33) | [0.911,2.230] | 1.038 (0.24) | [0.664,1.622] | 1.238 (0.29) | [0.787,1.950] |
| *Economic shocks* | | | | | | | | |
| Faced with new health expenses | 2.093*** (0.38) | [1.460,3.000] | 1.530*** (0.25) | [1.109,2.110] | 2.089*** (0.40) | [1.435,3.041] | 1.701*** (0.28) | [1.226,2.360] |
| Experienced adversity but did not reduce food consumption | 4.608** (3.59) | [1.003,21.173] | 1.475 (0.93) | [0.427,5.092] | 3.693* (2.66) | [0.899,15.172] | 1.615 (1.06) | [0.445,5.863] |
| Reduced food consumption as response to experienced adversity | 3.800 (3.22) | [0.721,20.038] | 1.249 (0.85) | [0.327,4.769] | 4.074* (3.27) | [0.845,19.648] | 2.556 (1.83) | [0.631,10.360] |
| *Educational disruption* | | | | | | | | |
| Enrolled in/Planning to enrol in full time education and not participating in learning activities | 0.559 (0.44) | [0.120,2.602] | 0.552 (0.33) | [0.173,1.763] | 1.190 (0.94) | [0.253,5.599] | 0.407 (0.28) | [0.107,1.546] |
| Enrolled in/Planning to enrol in full time education and participating in learning activities | 0.894 (0.16) | [0.625,1.278] | 1.127 (0.18) | [0.825,1.538] | 1.215 (0.23) | [0.841,1.754] | 0.898 (0.15) | [0.646,1.248] |
| *Past protective/risk factors* | | | | | | | | |
| Participant has long-term health problem, 2016 (Round 5) | 2.008*** (0.54) | [1.186,3.398] | 1.493* (0.33) | [0.964,2.312] | 1.910** (0.53) | [1.108,3.293] | 1.269 (0.28) | [0.827,1.947] |
| Total parent-child relationship score, 2012/2016 | 0.973 (0.03) | [0.922,1.028] | 0.977 (0.02) | [0.938,1.019] | 0.907*** | [0.856,0.961] | 0.931*** | [0.891,0.973] |

| | At least mild anxiety | | | | At least mild depression | | | |
|---|---|---|---|---|---|---|---|---|
| | Male | | Female | | Male | | Female | |
| | Odds Ratio | 95% CI | Odds Ratio | 95% CI | Odds Ratio | 95% CI | Odds Ratio | 95% CI |
| (Round 4/5) | | | | | (0.03) | | (0.02) | |
| Total peer-child relationship score, 2016 (Round 5) | 1.014 (0.03) | [0.954,1.078] | 0.989 (0.02) | [0.947,1.034] | 1.034 (0.03) | [0.968,1.104] | 1.023 (0.02) | [0.975,1.072] |
| Middle/Top wealth tercile R5, 2016 (Round 5) | 0.892 (0.18) | [0.600,1.325] | 1.247 (0.24) | [0.857,1.816] | 1.034 (0.21) | [0.695,1.537] | 1.239 (0.26) | [0.821,1.871] |
| N | 745 | | 751 | | 745 | | 751 | |

*Note: Odds ratios are unadjusted odds ratios. Robust standard errors in parenthesis, *** significant at 1%, ** significant at 5%, * significant at 10%. Base categories are as follows: Rural, Believe they are at no/low risk, Did not leave the house at all during the past 7 days, Did not spend more time taking care of children, Did not spend more time on household chores, Did not spend more time working in the family business, Did not face new health expenses, Did not suffer a shock, Never attended school or not enrolled in full-time education/not planning to enrol, Does not have long-term health condition, Lowest wealth tercile. All time-variant variables are measured in 2020 unless otherwise specified.*

### Supplementary Table 23. Logistic regression results: Symptoms of at least mild anxiety/depression by gender for the Younger Cohort only (Vietnam)

| | At least mild anxiety | | | | At least mild depression | | | |
|---|---|---|---|---|---|---|---|---|
| | Male | | Female | | Male | | Female | |
| | Odds Ratio | 95% CI | Odds Ratio | 95% CI | Odds Ratio | 95% CI | Odds Ratio | 95% CI |
| *Structural factors* | | | | | | | | |
| Age in months | 1.009 (0.04) | [0.933,1.091] | 1.027 (0.04) | [0.954,1.106] | 1.051 (0.04) | [0.976,1.131] | 1.014 (0.04) | [0.938,1.095] |
| Urban | 1.026 (0.30) | [0.576,1.826] | 1.550* (0.41) | [0.921,2.609] | 0.735 (0.24) | [0.386,1.399] | 1.562* (0.40) | [0.942,2.590] |
| *COVID-19 related changes/behaviours* | | | | | | | | |
| Risk perception: believe they are at medium/high risk | 1.553 (0.49) | [0.834,2.890] | 0.830 (0.25) | [0.464,1.482] | 1.711 (0.61) | [0.851,3.439] | 0.775 (0.22) | [0.446,1.347] |
| Left house for any reason in the past 7 days | 0.862 (0.33) | [0.409,1.814] | 1.783 (0.64) | [0.879,3.617] | 0.944 (0.37) | [0.441,2.022] | 1.480 (0.47) | [0.793,2.759] |
| Difference in subjective well-being between Round 5 (2016) and phone survey (2020) | 1.247*** (0.10) | [1.063,1.462] | 1.154** (0.07) | [1.021,1.305] | 1.444*** (0.12) | [1.221,1.708] | 1.008 (0.06) | [0.896,1.133] |
| *Change in responsibilities:* | | | | | | | | |
| Spend more time taking care of children | 1.005 (0.41) | [0.452,2.238] | 1.720** (0.46) | [1.019,2.903] | 1.425 (0.55) | [0.667,3.045] | 1.796** (0.44) | [1.111,2.902] |
| Spend more time on household chores | 0.948 (0.25) | [0.570,1.575] | 1.057 (0.26) | [0.654,1.706] | 1.079 (0.31) | [0.619,1.880] | 2.902*** (0.80) | [1.694,4.972] |

| | At least mild anxiety | | | | At least mild depression | | | |
|---|---|---|---|---|---|---|---|---|
| | Male | | Female | | Male | | Female | |
| | **Odds Ratio** | **95% CI** | **Odds Ratio** | **95% CI** | **Odds Ratio** | **95% CI** | **Odds Ratio** | **95% CI** |
| Spend more time working in the family business | 2.053** (0.73) | [1.018,4.140] | 1.509 (0.46) | [0.830,2.745] | 2.214** (0.79) | [1.099,4.459] | 1.750* (0.50) | [0.998,3.068] |
| *Economic shocks* | | | | | | | | |
| Faced with new health expenses | 0.930 (0.50) | [0.323,2.684] | 1.523 (0.62) | [0.688,3.368] | 1.294 (0.63) | [0.498,3.365] | 0.476 (0.24) | [0.181,1.255] |
| Experienced adversity but did not reduce food consumption | 2.415*** (0.77) | [1.295,4.502] | 1.091 (0.29) | [0.652,1.826] | 2.411*** (0.81) | [1.245,4.668] | 1.502 (0.40) | [0.888,2.542] |
| Reduced food consumption as response to experienced adversity | 2.963*** (1.22) | [1.320,6.652] | 1.166 (0.37) | [0.628,2.165] | 2.427** (1.04) | [1.050,5.613] | 2.449*** (0.74) | [1.350,4.441] |
| *Educational disruption* | | | | | | | | |
| Enrolled in/Planning to enrol in full time education and not participating in learning activities | 0.954 (0.44) | [0.390,2.333] | 0.817 (0.34) | [0.358,1.862] | 1.153 (0.58) | [0.431,3.085] | 0.967 (0.35) | [0.476,1.966] |
| Enrolled in/Planning to enrol in full time education and participating in learning activities | 0.678 (0.23) | [0.350,1.315] | 0.714 (0.19) | [0.420,1.216] | 1.250 (0.43) | [0.639,2.445] | 1.058 (0.29) | [0.622,1.799] |
| *Past protective/risk factors* | | | | | | | | |
| Participant has long-term health problem, 2016 (Round 5) | 1.190 (0.58) | [0.458,3.088] | 1.036 (0.37) | [0.517,2.074] | 2.696** (1.17) | [1.151,6.317] | 1.204 (0.40) | [0.630,2.300] |
| Total parent-child relationship score, 2012/2016 (Round 4/5) | 1.004 (0.05) | [0.918,1.098] | 1.055 (0.04) | [0.981,1.135] | 0.925 (0.05) | [0.833,1.026] | 0.999 (0.04) | [0.931,1.072] |
| Total peer-child relationship score, 2016 (Round 5) | 1.058 (0.06) | [0.950,1.178] | 0.916 (0.05) | [0.821,1.022] | 1.063 (0.07) | [0.926,1.220] | 0.948 (0.05) | [0.852,1.054] |
| Middle/Top wealth tercile R5, 2016 (Round 5) | 0.583* (0.18) | [0.323,1.055] | 0.741 (0.18) | [0.454,1.208] | 1.314 (0.45) | [0.672,2.566] | 1.164 (0.31) | [0.693,1.955] |
| N | 790 | | 806 | | 790 | | 806 | |

*Note: Odds ratios are unadjusted odds ratios. Robust standard errors in parenthesis, *** significant at 1%, ** significant at 5%, * significant at 10%. Base categories are as follows: Rural, Believe they are at no/low risk, Did not leave the house at all during the past 7 days, Did not spend more time taking care of children, Did not spend more time on household chores, Did not spend more time working in the family business, Did not face new health expenses, Did not suffer a shock, Never attended school or not enrolled in full-time education/not planning to enrol, Does not have long-term health condition, Lowest wealth tercile. All time-variant variables are measured in 2020 unless otherwise specified.*

**Supplementary Table 24. Ethiopia: Mean score and rates of general anxiety disorder**

| | Mean GAD-7 score | CI95% | p-value | Minimal (0-4) | CI95% | p-value | Mild anxiety (5-9) | CI95% | p-value | Moderate anxiety (10-14) | CI95% | p-value | Severe anxiety (>=15) | CI95% | p-value |
|---|---|---|---|---|---|---|---|---|---|---|---|---|---|---|---|
| **Total** | **2.06** | **1.94; 2.19** | | **82.13** | **80.46; 83.72** | | **15.21** | **13.73; 16.78** | | **2.34** | **1.74; 3.06** | | **0.32** | **0.13; 0.66** | |
| Male | 2.03 | 1.86; 2.20 | 0.603 | 82.52 | 80.35; 84.70 | 0.610 | 14.66 | 12.64; 16.69 | 0.445 | 2.47 | 1.58; 3.36 | 0.650 | 0.34 | 0.01; 0.67 | 0.856 |
| Female | 2.10 | 1.92; 2.28 | | 81.68 | 79.30; 84.07 | | 15.84 | 13.59; 18.10 | | 2.18 | 1.28; 3.08 | | 0.30 | -0.04; 0.63 | |
| Rural | 1.78 | 1.62; 1.93 | 0.000 | 85.33 | 83.30; 87.35 | 0.000 | 12.55 | 10.66; 14.45 | 0.000 | 2.12 | 1.30; 2.94 | 0.470 | 0.00 | 0.00; 0.00 | 0.004 |
| Urban | 2.40 | 2.20; 2.60 | | 78.39 | 75.84; 80.94 | | 18.33 | 15.93; 20.72 | | 2.59 | 1.61; 3.57 | | 0.70 | 0.18; 1.21 | |
| Poorest tercile | 1.89 | 1.68; 2.10 | 0.056 | 85.10 | 82.49; 87.71 | 0.011 | 11.98 | 9.60; 14.36 | 0.003 | 2.65 | 1.47; 3.82 | 0.502 | 0.28 | -0.11; 0.66 | 0.808 |
| Middle/Richest terciles | 2.15 | 1.99; 2.30 | | 80.68 | 78.66; 82.71 | | 16.79 | 14.88; 18.71 | | 2.18 | 1.44; 2.93 | | 0.34 | 0.04; 0.64 | |
| No internet | 1.90 | 1.74; 2.07 | 0.010 | 84.03 | 81.89; 86.18 | 0.017 | 13.20 | 11.22; 15.19 | 0.007 | 2.50 | 1.58; 3.41 | 0.608 | 0.27 | -0.04; 0.57 | 0.653 |
| With internet | 2.23 | 2.05; 2.41 | | 80.13 | 77.73; 82.53 | | 17.33 | 15.05; 19.60 | | 2.17 | 1.29; 3.04 | | 0.38 | 0.01; 0.75 | |
| N | 2183 | | | | | | | | | | | | | | |

*Notes: If any missing answers to questions then the whole score is set to missing. p-values represent significance of t-test of equality between groups (Male-Female; Rural-Urban; Bottom-Top/Middle wealth tercile; Internet access through home computer/working smartphone (No-Yes)). Poorest and middle/richest terciles refer to the household's position in the 2016 (round 5) wealth distribution.*
*Results are for the combined Younger Cohort/Older Cohort sample.*

### Supplementary Table 25. Ethiopia: Mean score and rates of depression

| | Mean PHQ-8 score | CI95% | p-val | PHQ (0-4) | CI95% | p-val | PHQ (5-9) | CI95% | p-val | PHQ (10-14) | CI95% | p-val | PHQ (15-19) | CI95% | p-val | PHQ (>=20) | CI95% | p-val |
|---|---|---|---|---|---|---|---|---|---|---|---|---|---|---|---|---|---|---|
| **Total** | **1.89** | **1.78; 2.01** | | **84.56** | **82.98; 86.05** | | **13.33** | **11.93; 14.83** | | **1.97** | **1.43; 2.64** | | **0.09** | **0.01; 0.33** | | **0.05** | **0.00; 0.25** | |
| Male | 1.86 | 1.70; 2.02 | 0.569 | 85.08 | 83.04; 87.12 | 0.470 | 12.79 | 10.87; 14.70 | 0.422 | 1.88 | 1.10; 2.65 | 0.733 | 0.17 | -0.07; 0.41 | 0.189 | 0.09 | -0.08; 0.25 | 0.354 |
| Female | 1.93 | 1.76; 2.10 | | 83.96 | 81.69; 86.23 | | 13.96 | 11.82; 16.10 | | 2.08 | 1.20; 2.96 | | 0.00 | 0.00; 0.00 | | 0.00 | 0.00; 0.00 | |
| Rural | 1.66 | 1.52; 1.81 | 0.000 | 87.11 | 85.19; 89.02 | 0.000 | 11.54 | 9.71; 13.36 | 0.007 | 1.27 | 0.63; 1.91 | 0.011 | 0.08 | -0.08; 0.25 | 0.909 | 0.00 | 0.00; 0.00 | 0.279 |
| Urban | 2.16 | 1.97; 2.34 | | 81.57 | 79.17; 83.97 | | 15.44 | 13.20; 17.68 | | 2.79 | 1.77; 3.81 | | 0.10 | -0.10; 0.29 | | 0.10 | -0.10; 0.29 | |
| Poorest tercile | 1.75 | 1.55; 1.95 | 0.086 | 85.65 | 83.09; 88.22 | 0.323 | 12.53 | 10.11; 14.96 | 0.444 | 1.53 | 0.63; 2.43 | 0.303 | 0.28 | -0.11; 0.66 | 0.043 | 0.00 | 0.00; 0.00 | 0.484 |
| Middle/Richest terciles | 1.96 | 1.82; 2.10 | | 84.03 | 82.15; 85.90 | | 13.72 | 11.96; 15.48 | | 2.18 | 1.44; 2.93 | | 0.00 | 0.00; 0.00 | | 0.07 | -0.07; 0.20 | |
| No internet | 1.69 | 1.54; 1.85 | 0.001 | 85.91 | 83.87; 87.94 | 0.074 | 12.49 | 10.55; 14.43 | 0.235 | 1.52 | 0.80; 2.23 | 0.118 | 0.09 | -0.09; 0.26 | 0.970 | 0.00 | 0.00; 0.00 | 0.304 |
| With internet | 2.10 | 1.93; 2.28 | | 83.15 | 80.89; 85.40 | | 14.22 | 12.12; 16.32 | | 2.45 | 1.52; 3.38 | | 0.09 | -0.09; 0.28 | | 0.09 | -0.09; 0.28 | |
| N | | 2183 | | | | | | | | | | | | | | | | |

*Notes: If any missing answers to questions then the whole score is set to missing. p-values represent significance of t-test of equality between groups (Male-Female; Rural-Urban; Bottom-Top/Middle wealth tercile; Internet access through home computer/working smartphone (No-Yes)). Poorest and middle/richest terciles refer to the household's position in the 2016 (round 5) wealth distribution.*
*Results are for the combined Younger Cohort/ Older Cohort sample.*

### Supplementary Table 26.India: Mean score and rates of general anxiety disorder

| | Mean GAD-7 score | CI95% | p-value | Minimal (0-4) | CI95% | p-value | Mild anxiety (5-9) | CI95% | p-value | Moderate anxiety (10-14) | CI95% | p-value | Severe anxiety (>=15) | CI95% | p-value |
|---|---|---|---|---|---|---|---|---|---|---|---|---|---|---|---|
| **Total** | **1.69** | **1.59; 1.78** | | **88.94** | **87.68; 90.12** | | **9.15** | **8.08; 10.32** | | **1.64** | **1.19; 2.20** | | **0.27** | **0.11; 0.55** | |
| Male | 1.55 | 1.43; 1.67 | 0.003 | 90.72 | 89.19; 92.26 | 0.002 | 8.04 | 6.59; 9.48 | 0.038 | 1.10 | 0.54; 1.65 | 0.022 | 0.15 | -0.06; 0.35 | 0.210 |
| Female | 1.84 | 1.69; 1.99 | | 86.99 | 85.13; 88.86 | | 10.38 | 8.69; 12.06 | | 2.23 | 1.42; 3.05 | | 0.40 | 0.05; 0.75 | |
| Rural | 1.76 | 1.65; 1.88 | 0.014 | 88.19 | 86.73; 89.66 | 0.055 | 9.56 | 8.23; 10.90 | 0.252 | 1.92 | 1.30; 2.55 | 0.071 | 0.32 | 0.06; 0.58 | 0.401 |
| Urban | 1.50 | 1.34; 1.66 | | 90.80 | 88.73; 92.87 | | 8.13 | 6.17; 10.09 | | 0.93 | 0.24; 1.62 | | 0.13 | -0.13; 0.39 | |
| Poorest tercile | 1.83 | 1.66; 2.00 | 0.042 | 86.48 | 84.22; 88.74 | 0.004 | 11.02 | 8.95; 13.09 | 0.018 | 2.16 | 1.20; 3.12 | 0.137 | 0.34 | -0.04; 0.73 | 0.602 |
| Middle/Richest terciles | 1.62 | 1.50; 1.73 | | 90.18 | 88.79; 91.58 | | 8.21 | 6.92; 9.50 | | 1.38 | 0.83; 1.93 | | 0.23 | 0.00; 0.45 | |
| No internet | 1.80 | 1.52; 2.08 | 0.410 | 87.84 | 84.11; 91.57 | 0.521 | 10.47 | 6.98; 13.97 | 0.403 | 1.69 | 0.22; 3.16 | 0.944 | 0.00 | 0.00; 0.00 | 0.345 |
| With internet | 1.67 | 1.57; 1.78 | | 89.08 | 87.81; 90.35 | | 8.99 | 7.82; 10.15 | | 1.63 | 1.12; 2.15 | | 0.30 | 0.08; 0.52 | |
| N | 2622 | | | | | | | | | | | | | | |

*Notes: If any missing answers to questions then the whole score is set to missing. p-values represent significance of t-test of equality between groups (Male-Female; Rural-Urban; Bottom-Top/Middle wealth tercile; Internet access through home computer/working smartphone (No-Yes)). Poorest and middle/richest terciles refer to the household's position in the 2016 (round 5) wealth distribution.*
*Results are for the combined Younger Cohort/ Older Cohort sample.*

**Supplementary Table 27. India: Mean score and rates of depression**

| | Mean PHQ-8 score | CI95% | p-value | No significant depressive symptoms(0-4) | CI95% | p-value | Mild depression (5-9) | CI95% | p-value | Moderate depressive symptoms(10-14) | CI95% | p-value | Moderately severe depressive symptoms(15-19) | CI95% | p-value |
|---|---|---|---|---|---|---|---|---|---|---|---|---|---|---|---|
| **Total** | **1.41** | **1.33; 1.5** | | **90.08** | **88.88; 91.2** | | **8.73** | **7.68; 9.88** | | **1.03** | **0.68; 1.49** | | **0.15** | **0.04; 0.39** | |
| Male | 1.34 | 1.22; 1.45 | 0.067 | 90.36 | 88.79; 91.92 | 0.624 | 8.84 | 7.33; 10.34 | 0.843 | 0.73 | 0.28; 1.18 | 0.113 | 0.07 | -0.07; 0.22 | 0.276 |
| Female | 1.50 | 1.37; 1.63 | | 89.78 | 88.11; 91.46 | | 8.62 | 7.06; 10.17 | | 1.36 | 0.72; 2.00 | | 0.24 | -0.03; 0.51 | |
| Rural | 1.47 | 1.36; 1.58 | 0.039 | 89.16 | 87.75; 90.57 | 0.012 | 9.40 | 8.08; 10.72 | 0.056 | 1.28 | 0.77; 1.79 | 0.043 | 0.16 | -0.02; 0.34 | 0.873 |
| Urban | 1.27 | 1.12; 1.42 | | 92.40 | 90.50; 94.30 | | 7.07 | 5.23; 8.90 | | 0.40 | -0.05; 0.85 | | 0.13 | -0.13; 0.39 | |
| Poorest tercile | 1.49 | 1.33; 1.65 | 0.220 | 89.20 | 87.15; 91.26 | 0.284 | 9.55 | 7.60; 11.49 | 0.296 | 1.02 | 0.36; 1.69 | 0.980 | 0.23 | -0.09; 0.54 | 0.486 |
| Middle/Richest terciles | 1.37 | 1.27; 1.48 | | 90.53 | 89.15; 91.90 | | 8.32 | 7.03; 9.620 | | 1.03 | 0.56; 1.51 | | 0.11 | -0.04; 0.27 | |
| No internet | 1.42 | 1.17; 1.67 | 0.941 | 89.86 | 86.42; 93.31 | 0.894 | 8.78 | 5.55; 12.02 | 0.974 | 1.35 | 0.03; 2.67 | 0.561 | 0.00 | 0.00; 0.00 | 0.475 |
| With internet | 1.41 | 1.32; 1.50 | | 90.11 | 88.90; 91.33 | | 8.73 | 7.58; 9.88 | | 0.99 | 0.59; 1.39 | | 0.17 | 0.00; 0.34 | |
| N | 2622 | | | | | | | | | | | | | | |

*Notes: If any missing answers to questions then the whole score is set to missing. p-values represent significance of t-test of equality between groups (Male-Female; Rural-Urban; Bottom-Top/Middle wealth tercile; Internet access through home computer/working smartphone (No-Yes)). Poorest and middle/richest terciles refer to the household's position in the 2016 (round 5) wealth distribution.*
*Results are for the combined Younger Cohort/ Older Cohort sample. No severe depressive symptoms were reported.*

### Supplementary Table 28. Peru: Mean score and rates of general anxiety disorder

| | Mean GAD-7 score | CI95% | p-value | Minimal (0-4) | CI95% | p-value | Mild anxiety (5-9) | CI95% | p-value | Moderate anxiety (10-14) | CI95% | p-value | Severe anxiety (>=15) | CI95% | p-value |
|---|---|---|---|---|---|---|---|---|---|---|---|---|---|---|---|
| **Total** | **4.53** | **4.34; 4.72** | | **59.14** | **56.88; 61.37** | | **27.34** | **25.34; 29.42** | | **10.55** | **9.20; 12.02** | | **2.97** | **2.25; 3.84** | |
| Male | 3.80 | 3.55; 4.04 | 0.000 | 66.35 | 63.36; 69.35 | 0.000 | 23.72 | 21.02; 26.42 | 0.000 | 7.94 | 6.23; 9.66 | 0.000 | 1.99 | 1.10; 2.87 | 0.011 |
| Female | 5.29 | 5.01; 5.57 | | 51.72 | 48.51; 54.94 | | 31.08 | 28.10; 34.05 | | 13.23 | 11.05; 15.41 | | 3.98 | 2.72; 5.24 | |
| Rural | 3.87 | 3.46; 4.27 | 0.001 | 66.29 | 61.37; 71.21 | 0.002 | 25.56 | 21.02; 30.10 | 0.402 | 5.62 | 3.22; 8.01 | 0.001 | 2.53 | 0.89; 4.16 | 0.588 |
| Urban | 4.69 | 4.47; 4.90 | | 57.48 | 55.00; 59.96 | | 27.76 | 25.51; 30.00 | | 11.69 | 10.08; 13.30 | | 3.07 | 2.20; 3.93 | |
| Poorest tercile | 4.05 | 3.74; 4.35 | 0.001 | 63.08 | 59.16; 66.99 | 0.020 | 27.01 | 23.41; 30.61 | 0.826 | 8.21 | 5.98; 10.43 | 0.026 | 1.71 | 0.66; 2.76 | 0.031 |
| Middle/Richest terciles | 4.75 | 4.51; 4.99 | | 57.37 | 54.68; 60.06 | | 27.50 | 25.07; 29.92 | | 11.60 | 9.86; 13.34 | | 3.53 | 2.53; 4.54 | |
| No internet | 4.96 | 4.14; 5.78 | 0.367 | 48.68 | 37.37; 60.00 | 0.058 | 36.84 | 25.92; 47.77 | 0.058 | 14.47 | 6.51; 22.44 | 0.255 | 0.00 | 0.00; 0.00 | 0.120 |
| With internet | 4.51 | 4.32; 4.71 | | 59.58 | 57.32; 61.84 | | 26.95 | 24.90; 28.99 | | 10.38 | 8.97; 11.79 | | 3.09 | 2.29; 3.89 | |
| N | 1887 | | | | | | | | | | | | | | |

*Notes: If any missing answers to questions then the whole score is set to missing. p-values represent significance of t-test of equality between groups (Male-Female; Rural-Urban; Bottom-Top/Middle wealth tercile; Internet access through home computer/working smartphone (No-Yes)). Poorest and middle/richest terciles refer to the household's position in the 2016 (round 5) wealth distribution.*
*Results are for the combined Younger Cohort/ Older Cohort sample.*

### Supplementary Table 29. Peru: Mean score and rates of depression

| | Mean PHQ-8 score | CI 95% | p-val | PHQ (0-4) | CI 95% | p-val | PHQ (5-9) | CI 95% | p-val | PHQ (10-14) | CI 95% | p-val | PHQ (15-19) | CI 95% | p-val | PHQ (>=20) | CI 95% | p-val |
|---|---|---|---|---|---|---|---|---|---|---|---|---|---|---|---|---|---|---|
| **Total** | **3.61** | **3.43; 3.79** | | **68.42** | **66.26; 70.51** | | **21.94** | **20.09; 23.88** | | **7.79** | **6.62; 9.09** | | **1.48** | **0.99; 2.14** | | **0.37** | **0.15; 0.76** | |
| Male | 3.08 | 2.85; 3.32 | 0.000 | 72.62 | 69.79; 75.45 | 0.000 | 19.75 | 17.22; 22.27 | 0.020 | 6.58 | 5.01; 8.16 | 0.047 | 0.73 | 0.19; 1.27 | 0.006 | 0.31 | -0.04; 0.67 | 0.677 |
| Female | 4.15 | 3.89; 4.41 | | 64.09 | 61.00; 67.17 | | 24.19 | 21.44; 26.95 | | 9.03 | 7.19; 10.88 | | 2.26 | 1.30; 3.21 | | 0.43 | 0.01; 0.85 | |
| Rural | 3.11 | 2.73; 3.49 | 0.008 | 73.60 | 69.01; 78.18 | 0.020 | 19.38 | 15.27; 23.50 | 0.196 | 6.18 | 3.67; 8.69 | 0.208 | 0.84 | -0.11; 1.79 | 0.267 | 0.00 | 0.00; 0.00 | 0.201 |
| Urban | 3.73 | 3.53; 3.92 | | 67.21 | 64.86; 69.56 | | 22.53 | 20.44; 24.63 | | 8.16 | 6.79; 9.54 | | 1.63 | 1.00; 2.27 | | 0.46 | 0.12; 0.80 | |
| Poorest tercile | 3.07 | 2.79; 3.36 | 0.000 | 73.85 | 70.28; 77.41 | 0.001 | 19.66 | 16.43; 22.88 | 0.109 | 5.47 | 3.62; 7.32 | 0.012 | 0.68 | 0.01; 1.35 | 0.054 | 0.34 | -0.13; 0.82 | 0.889 |
| Middle/ Richest terciles | 3.85 | 3.63; 4.07 | | 65.98 | 63.40; 68.55 | | 22.96 | 20.68; 25.25 | | 8.83 | 7.29; 10.38 | | 1.84 | 1.11; 2.57 | | 0.38 | 0.05; 0.72 | |
| No internet | 3.34 | 2.62; 4.07 | 0.545 | 68.42 | 57.89; 78.95 | 0.999 | 25.00 | 15.19; 34.81 | 0.511 | 6.58 | 0.96; 12.19 | 0.688 | 0.00 | 0.00; 0.00 | 0.275 | 0.00 | 0.00; 0.00 | 0.587 |
| With internet | 3.62 | 3.44; 3.80 | | 68.42 | 66.27; 70.56 | | 21.81 | 19.91; 23.71 | | 7.84 | 6.60; 9.08 | | 1.55 | 0.98; 2.11 | | 0.39 | 0.10; 0.67 | |
| N | 1887 | | | | | | | | | | | | | | | | | |

*Notes: If any missing answers to questions then the whole score is set to missing. p-values represent significance of t-test of equality between groups (Male-Female; Rural-Urban; Bottom-Top/Middle wealth tercile; Internet access through home computer/working smartphone (No-Yes)). Poorest and middle/richest terciles refer to the household's position in the 2016 (round 5) wealth distribution.*
*Results are for the combined Younger Cohort/ Older Cohort sample.*

### Supplementary Table 30. Vietnam: Mean score and rates of general anxiety disorder

| | Mean GAD-7 score | CI95% | p-value | Minimal (0-4) | CI95% | p-value | Mild anxiety (5-9) | CI95% | p-value | Moderate anxiety (10-14) | CI95% | p-value | Severe anxiety (>=15) | CI95% | p-value |
|---|---|---|---|---|---|---|---|---|---|---|---|---|---|---|---|
| **Total** | **1.47** | **1.37; 1.56** | | **90.68** | **89.42; 91.84** | | **7.93** | **6.85; 9.11** | | **1.09** | **0.71; 1.60** | | **0.30** | **0.12; 0.63** | |
| Male | 1.31 | 1.17; 1.44 | 0.001 | 92.08 | 90.49; 93.67 | 0.025 | 6.48 | 5.03; 7.93 | 0.013 | 1.17 | 0.54; 1.80 | 0.717 | 0.27 | -0.04; 0.58 | 0.769 |
| Female | 1.62 | 1.48; 1.76 | | 89.37 | 87.61; 91.12 | | 9.28 | 7.63; 10.94 | | 1.01 | 0.44; 1.58 | | 0.34 | 0.01; 0.67 | |
| Rural | 1.47 | 1.34; 1.60 | 0.925 | 90.93 | 89.33; 92.53 | 0.652 | 7.62 | 6.15; 9.10 | 0.559 | 1.04 | 0.48; 1.61 | 0.819 | 0.40 | 0.05; 0.75 | 0.362 |
| Urban | 1.46 | 1.32; 1.60 | | 90.38 | 88.60; 92.17 | | 8.29 | 6.62; 9.95 | | 1.14 | 0.50; 1.79 | | 0.19 | -0.07; 0.45 | |
| Poorest tercile | 1.60 | 1.42; 1.78 | 0.045 | 88.33 | 86.08; 90.59 | 0.006 | 10.00 | 7.89; 12.11 | 0.008 | 1.15 | 0.40; 1.90 | 0.830 | 0.51 | 0.01; 1.01 | 0.195 |
| Middle/Richest terciles | 1.40 | 1.29; 1.51 | | 91.89 | 90.51; 93.26 | | 6.86 | 5.59; 8.13 | | 1.06 | 0.54; 1.57 | | 0.20 | -0.03; 0.42 | |
| No internet | 2.40 | 1.24; 3.56 | 0.017 | 77.14 | 63.02; 91.26 | 0.005 | 17.14 | 4.47; 29.82 | 0.042 | 2.86 | -2.75; 8.46 | 0.310 | 2.86 | -2.75; 8.46 | 0.006 |
| With internet | 1.45 | 1.36; 1.55 | | 90.89 | 89.70; 92.08 | | 7.78 | 6.68; 8.89 | | 1.06 | 0.64; 1.48 | | 0.27 | 0.05; 0.48 | |
| N | 2296 | | | | | | | | | | | | | | |

*Notes: If any missing answers to questions then the whole score is set to missing. p-values represent significance of t-test of equality between groups (Male-Female; Rural-Urban; Bottom-Top/Middle wealth tercile; Internet access through home computer/working smartphone (No-Yes)). Poorest and middle/richest terciles refer to the household's position in the 2016 (round 5) wealth distribution.*
*Results are for the combined Younger Cohort/ Older Cohort sample.*

### Supplementary Table 31. Vietnam: Mean score and rates of depression

| | Mean PHQ-8 score | CI95% | p-value | No significant depressive symptoms(0-4) | CI95% | p-value | Mild depression (5-9) | CI95% | p-value | Moderate depr-essive symptoms (10-14) | CI95% | p-value | Moderately severe depressive symptoms(15-19) | CI95% | p-value |
|---|---|---|---|---|---|---|---|---|---|---|---|---|---|---|---|
| **Total** | **1.42** | **1.32; 1.52** | | **90.51** | **89.23; 91.67** | | **7.67** | **6.61; 8.83** | | **1.39** | **0.96; 1.96** | | **0.44** | **0.21; 0.80** | |
| Male | 1.24 | 1.11; 1.37 | 0.000 | 92.35 | 90.78; 93.91 | 0.004 | 6.12 | 4.71; 7.53 | 0.007 | 1.17 | 0.54; 1.80 | 0.376 | 0.36 | 0.01; 0.71 | 0.595 |
| Female | 1.59 | 1.45; 1.74 | | 88.78 | 86.98; 90.58 | | 9.11 | 7.47; 10.75 | | 1.60 | 0.89; 2.32 | | 0.51 | 0.10; 0.91 | |
| Rural | 1.43 | 1.30; 1.56 | 0.852 | 90.61 | 88.99; 92.23 | 0.852 | 7.70 | 6.22; 9.19 | 0.939 | 1.20 | 0.6; 1.81 | 0.398 | 0.48 | 0.10; 0.87 | 0.716 |
| Urban | 1.41 | 1.26; 1.56 | | 90.38 | 88.60; 92.17 | | 7.62 | 6.01; 9.23 | | 1.62 | 0.85; 2.38 | | 0.38 | 0.01; 0.75 | |
| Poorest tercile | 1.40 | 1.23; 1.57 | 0.757 | 91.54 | 89.58; 93.49 | 0.226 | 6.28 | 4.58; 7.99 | 0.074 | 1.54 | 0.67; 2.40 | 0.671 | 0.64 | 0.08; 1.20 | 0.284 |
| Middle/Richest terciles | 1.43 | 1.31; 1.55 | | 89.97 | 88.46; 91.49 | | 8.38 | 6.98; 9.77 | | 1.32 | 0.74; 1.89 | | 0.33 | 0.04; 0.62 | |
| No internet | 2.20 | 1.05; 3.35 | 0.053 | 85.71 | 73.95; 97.48 | 0.330 | 8.57 | -0.84; 17.99 | 0.839 | 2.86 | -2.75; 8.46 | 0.457 | 2.86 | -2.75; 8.46 | 0.028 |
| With internet | 1.41 | 1.31; 1.51 | | 90.58 | 89.37; 91.78 | | 7.65 | 6.55; 8.75 | | 1.37 | 0.89; 1.85 | | 0.40 | 0.14; 0.66 | |
| N | 2296 | | | | | | | | | | | | | | |

*Notes: If any missing answers to questions then the whole score is set to missing. p-values represent significance of t-test of equality between groups (Male-Female; Rural-Urban; Bottom-Top/Middle wealth tercile; Internet access through home computer/working smartphone (No-Yes)). Poorest and middle/richest terciles refer to the household's position in the 2016 (round 5) wealth distribution.*
*Results are for the combined Younger Cohort/ Older Cohort sample. No severe depressive symptoms were reported.*

### Supplementary Table 32. Correlation between GAD-7 and PHQ-8 scores

| | Ethiopia | India | Peru | Vietnam |
|---|---|---|---|---|
| Correlation coefficient | 0.629*** | 0.610*** | 0.700*** | 0.636*** |

*Note: Bonferri corrected p-value *** significant at 1%, ** significant at 5%, * significant at 10% for the unconditional correlation between GAD-7 and PHQ-8. Results are for the combined Younger Cohort/ Older Cohort sample.*

### Supplementary Table 33. Joint rates

| | Ethiopia | | India | | Peru | | Vietnam | |
|---|---|---|---|---|---|---|---|---|
| | % of sample | 95% CI | % of sample | 95% CI | % of sample | 95% CI | % of sample | 95% CI |
| Display neither symptoms of anxiety nor depression | 75.45 | 73.58; 77.24 | 83.87 | 82.40; 85.26 | 52.36 | 50.08; 54.63 | 85.50 | 83.99; 86.91 |
| Display symptoms of at least mild anxiety only | 9.12 | 7.94; 10.4 | 6.22 | 5.32; 7.21 | 16.06 | 14.43; 17.79 | 5.01 | 4.15; 5.98 |
| Display symptoms of at least mild depression only | 6.69 | 7.94; 10.4 | 5.07 | 5.32; 7.21 | 6.78 | 14.43; 17.79 | 5.18 | 4.15; 5.98 |
| Display symptoms of at least mild anxiety and at least mild depression | 8.75 | 7.60; 10.01 | 4.84 | 4.05; 5.74 | 24.80 | 22.87; 26.81 | 4.31 | 3.52; 5.22 |

*Note: Results are for the combined Younger Cohort/ Older Cohort sample.*

**Supplementary Table 34. Correlation between subjective well-being and GAD-7/PHQ-8 scores**

| | Ethiopia | | | | India | | | | Peru | | | | Vietnam | | | |
|---|---|---|---|---|---|---|---|---|---|---|---|---|---|---|---|---|
| | Subjective well-being (phone survey, 2020) | | Subjective well-being (difference Round 5 (2016) and phone survey (2020)) | | Subjective well-being (phone survey, 2020) | | Subjective well-being (difference Round 5 (2016) and phone survey (2020)) | | Subjective well-being (phone survey, 2020) | | Subjective well-being (difference Round 5 (2016) and phone survey (2020)) | | Subjective well-being (phone survey, 2020) | | Subjective well-being (difference Round 5 (2016) and phone survey (2020)) | |
| | r | p-value | r | p-value | r | p-value | r | p-value | r | p-value | r | p-value | r | p-value | r | p-value |
| Mean GAD-7 score | -0.140 | 0.000 | 0.041 | 0.054 | -0.107 | 0.000 | 0.064 | 0.001 | -0.199 | 0.000 | 0.096 | 0.000 | -0.253 | 0.000 | 0.118 | 0.000 |
| Mean PHQ-8 score | -0.091 | 0.000 | 0.024 | 0.263 | -0.097 | 0.000 | 0.055 | 0.005 | -0.186 | 0.000 | 0.067 | 0.004 | -0.207 | 0.000 | 0.119 | 0.000 |

*Note: Results are for the combined Younger Cohort/ Older Cohort sample.*

**Supplementary Table 35. Subjective well-being score Round 5 (2016) / phone survey (2020) by anxiety/depression (Ethiopia & India)**

| | Ethiopia | | | | | | | India | | | | | | |
|---|---|---|---|---|---|---|---|---|---|---|---|---|---|---|
| | **All** | **at least mild anxiety** | **no anxiety** | **p-value** | **at least mild depression** | **no depression** | **p-value** | **All** | **at least mild anxiety** | **no anxiety** | **p-value** | **at least mild depression** | **no depression** | **p-value** |
| (1) Subjective well-being, 2016 (Round 5) | 5.69 | 5.43 | 5.75 | 0.000 | 5.61 | 5.71 | 0.303 | 5.05 | 4.94 | 5.06 | 0.178 | 4.98 | 5.06 | 0.381 |
| (2) Subjective well-being, 2020 (Phone survey) | 4.73 | 4.28 | 4.83 | 0.000 | 4.36 | 4.80 | 0.000 | 4.56 | 4.18 | 4.61 | 0.000 | 4.12 | 4.61 | 0.000 |
| Difference in subjective well-being (1)-(2) | 0.96 | 1.15 | 0.92 | 0.052 | 1.26 | 0.91 | 0.005 | 0.49 | 0.77 | 0.45 | 0.009 | 0.86 | 0.45 | 0.001 |

*Note: Results are for the combined Younger Cohort/ Older Cohort sample.*

**Supplementary Table 36. Subjective well-being score Round 5 (2016) / phone survey (2020) by anxiety/depression (Peru & Vietnam)**

| | Peru | | | | | | | Vietnam | | | | | | |
|---|---|---|---|---|---|---|---|---|---|---|---|---|---|---|
| | **All** | **at least mild anxiety** | **no anxiety** | **p-value** | **at least mild depression** | **no depression** | **p-value** | **All** | **at least mild anxiety** | **no anxiety** | **p-value** | **at least mild depression** | **no depression** | **p-value** |
| (1) Subjective well-being, 2016 (Round 5) | 6.33 | 6.20 | 6.42 | 0.001 | 6.14 | 6.42 | 0.000 | 5.87 | 5.46 | 5.91 | 0.000 | 5.65 | 5.89 | 0.024 |
| (2) Subjective well-being, 2020 (Phone survey) | 5.81 | 5.47 | 6.05 | 0.000 | 5.44 | 5.98 | 0.000 | 6.26 | 5.33 | 6.36 | 0.000 | 5.54 | 6.34 | 0.000 |
| Difference in subjective well-being (1)-(2) | 0.52 | 0.74 | 0.38 | 0.000 | 0.70 | 0.44 | 0.013 | -0.39 | 0.14 | -0.45 | 0.000 | 0.11 | -0.45 | 0.000 |

*Note: Results are for the combined Younger Cohort/ Older Cohort sample.*
